# Development and application of an evidence-based directed acyclic graph to evaluate the associations between metal mixtures and cardiometabolic outcomes

**DOI:** 10.1101/2021.03.05.21252993

**Authors:** Emily Riseberg, Rachel D. Melamed, Katherine A. James, Tanya L. Alderete, Laura Corlin

## Abstract

**Objectives:** Specifying analytic models to assess relationships among metal mixtures and cardiometabolic outcomes requires evidence-based models of the causal structures; however, such models have not been previously published. The objective of this study was to develop and evaluate a directed acyclic graph diagraming metal mixture exposure and cardiometabolic outcomes.

**Methods:** We conducted a systematic literature search to develop the directed acyclic graph (DAG) of metal mixtures and cardiometabolic outcomes. To evaluate consistency of the DAG, we tested the suggested conditional independence statements using linear and logistic regression analyses with data from the San Luis Valley Diabetes Study (SLVDS; n=1795). We compared the proportion of statements supported by the data to the proportion of conditional independence statements supported by 100 DAGs with the same structure but randomly permuted nodes. Next, we used our DAG to identify minimally sufficient adjustment sets needed to estimate the association between metal mixtures and cardiometabolic outcomes in the SLVDS and applied them using Bayesian kernel machine regression models.

**Results:** From the 42 articles included in the review, we developed an evidence-based DAG with 163 testable conditional independence statements (64% supported by SLVDS data). Only 5% of DAGs with randomly permuted nodes indicated more agreement with the data than our evidence-based DAG. We did not observe evidence for an association between metal mixtures and cardiometabolic outcomes in the pilot analysis.

**Conclusions:** We developed, tested, and applied an evidence-based approach to analyze associations between metal mixtures and cardiometabolic health.

## Introduction

Extensive epidemiologic and toxicologic evidence indicates that metals and metalloids [e.g., arsenic (As), cadmium (Cd), manganese (Mn), and tungsten (W); hereafter simplified as ‘metals’] are associated with cardiometabolic outcomes (Kuo et al. 2013; Tyrrell et al. 2013; Tellez-Plaza et al. 2013; Wang et al. 2014; Edwards and Ackerman 2016; Nigra et al. 2016; Shan Zhilei et al. 2016; Wu et al. 2016; K. A. Moon et al. 2017; Tinkov et al. 2018; Long et al. 2019; Riseberg et al. 2021). Whereas most of the published studies considered exposure to only one metal, numerous calls exist to examine the associations between metal mixtures and cardiometabolic outcomes (Tyrrell et al. 2013; Corlin et al. 2016; Li and Yang 2018). Examining the health outcomes associated with metal mixtures would more realistically reflect environmental exposure conditions (Cui et al. 2005; Pang et al. 2016). Yet even analyses that include multiple metals typically only adjust for concentrations of non-target metals (not accounting for interactions among metal mixtures) or use stratified analyses. These stratified analyses, often in the form of associations with low exposure to metal A/low exposure to metal B versus high exposure to metal A/high exposure to metal B, do not capture complex non-linear relationships (Tyrrell et al. 2013; Rajpathak et al. 2004; Mendy, Gasana, and Vieira 2012; Bobb et al. 2015; Tsai et al. 2017).

Development of analytic models probing the complex relationships among metal mixtures and cardiometabolic outcomes requires an understanding of the putative underlying causal structure. An evidence-based directed acyclic graph (DAG) is one way to represent such a causal structure. DAGs clarify causal contrasts and explicitly show assumptions about common causes of exposures and outcomes (e.g., sources of metal (co)exposure that also affect cardiometabolic health) that we need to account for in our study design and/or analysis (Pearl 1995; Tennant et al. 2020; Pearce and Lawlor 2016). They can additionally reveal sources of selection bias or collider bias (Arah 2019). DAGs are also useful for identifying minimally sufficient adjustment sets of variables; when DAGs are used in this way, articles should report the assumed DAG (Tennant et al. 2020). Few evidence-based DAGs exist in the environmental epidemiology context due to the need to conduct literature reviews and to empirically test the applicability of the DAG for the study context (as one example, see Corlin et al., 2016). No evidence-based DAGs have been previously published describing the structure underlying potential metal mixture-cardiometabolic outcome relationships. Such a DAG could help researchers assess how specific environmentally relevant metal mixtures mechanistically affect the development of cardiometabolic outcomes. Therefore, our primary objective was to conduct a systematic literature search to support the development of an evidence-based DAG diagraming the relationships among exposure to metal mixtures, the development of cardiometabolic outcomes, and potential common causes of exposures and outcomes. Our secondary objective was to evaluate this DAG and apply it to a real environmental health context using data from a cohort of adults residing in the rural San Luis Valley of Colorado.

## Materials and Methods

### Literature review and directed acyclic graph development

We conducted a systematic search in PubMed using the following text: ((tungsten[MeSH Terms]) OR (uranium[MeSH Terms]) OR (cadmium[MeSH Terms]) OR (arsenic[MeSH Terms]) OR (manganese[MeSH Terms])) AND ((cardiovascular disease[MeSH Terms]) OR (type 2 diabetes mellitus[MeSH Terms]) OR (hypertension[MeSH Terms])) AND ((“review”[Publication Type]) OR (“systematic review”[Publication Type]) OR (“meta analysis”[Publication Type])) AND (“2011/01/01”[Date - Publication] : “3000”[Date - Publication]).

The search strategy is detailed in **Figure 1**. We specified our search to reviews, systematic reviews, and meta-analyses published between the beginning of 2011 to the time of the search (2021) in English. Articles had to review human epidemiological studies and have one of the metals (As, Cd, Mn, W, and U) as an exposure and either cardiovascular disease, diabetes, or hypertension as an outcome. We additionally added five articles from the authors’ prior knowledge that fit the criteria of the search.

**Figure 1.**
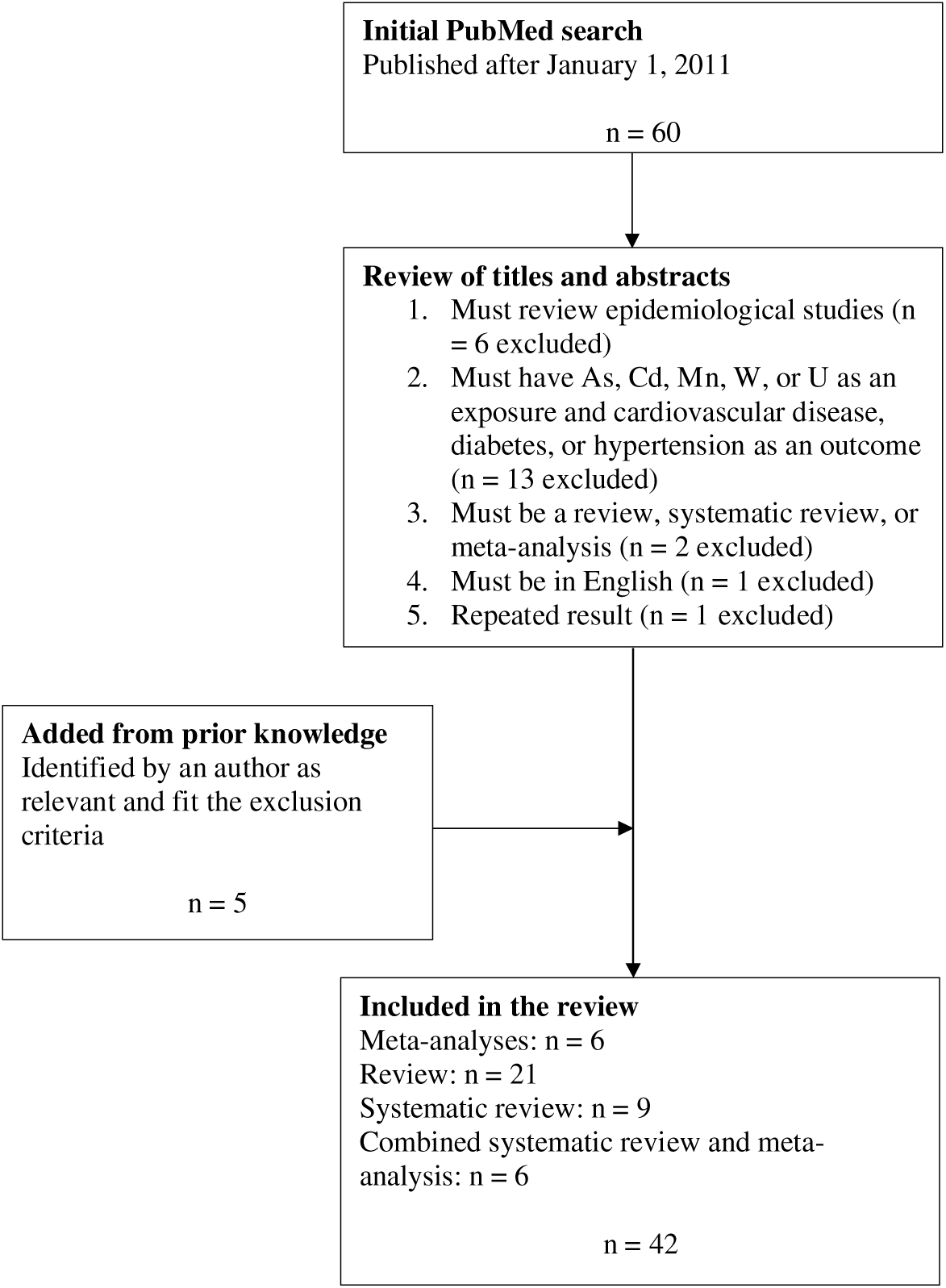
Literature review strategy

From each article included in the review, we collected the following information: authors, year, journal, publication type, study population, exposure, exposure sources, outcome, and main results. Based on the extracted data, we created DAGs using the software DAGitty (Textor et al. 2016). Each arrow from an exposure to an outcome represented a relationship mentioned in at least one articles from the literature review (**Figure 2**) (Xu, Mondal, and Polya 2020; Leng et al. 2019; da Cunha Martins et al. 2018; Chowdhury et al. 2018; Kuo et al. 2017; Beck, Styblo, and Sethupathy 2017; Khan et al. 2017; Phung et al. 2017; K. A. Moon et al. 2017; Alamolhodaei, Shirani, and Karimi 2015; Abdul et al. 2015; Ellinsworth 2015; Tsuji et al. 2014; Solenkova et al. 2014; Wang et al. 2014; Stea et al. 2014; K. Moon, Guallar, and Navas–Acien 2012; Kuo et al. 2013; Andra et al. 2013; Boekelheide et al. 2012; Abhyankar et al. 2012; Jomova et al. 2011; Diaz et al. 2021; Martins et al. 2021; Little et al. 2020; Tinkov et al. 2018; Satarug, Vesey, and Gobe 2017; Edwards and Ackerman 2016; Kukongviriyapan, Apaijit, and Kukongviriyapan 2016; Larsson and Wolk 2016; Hecht et al. 2013; Tellez-Plaza et al. 2013; Caciari et al. 2013; Thévenod and Lee 2013; Satarug and Moore 2012; Li and Yang 2018; Sanjeevi et al. 2018; Kaur and Henry 2014; Siddiqui, Bawazeer, and Scaria Joy 2014; Corlin et al. 2016; Nigra et al. 2016; Zhivin, Laurier, and Canu 2014). For the other variables in the DAG, we followed the following steps: (1) identified sources of exposure from the literature (i.e., diet, ambient air, smoking, soil, and drinking water), (2) identified risk factors for the outcomes using consensus statements (i.e., sex, age, obesity, education, ethnicity, income, physical activity, and alcohol intake), (Alberti, Zimmet, and Shaw 2007; Havranek et al. 2015; Whelton et al. 2018; Wood 2001) (3) conducted a search on PubMed for risk factors and outcomes if a risk factor for one outcome was not identified as a risk factor for another outcome (e.g., “smoking diabetes”), (4) identified risk factor—risk factor and source of exposure—risk factor associations through a PubMed search containing the two words (e.g., “education ethnicity” or “soil ethnicity”). The sources for each arrow are provided in **Appendix A**, and the DAG is shown in **Figure 2** (code provided in **Appendix B**).

**Figure 2.**
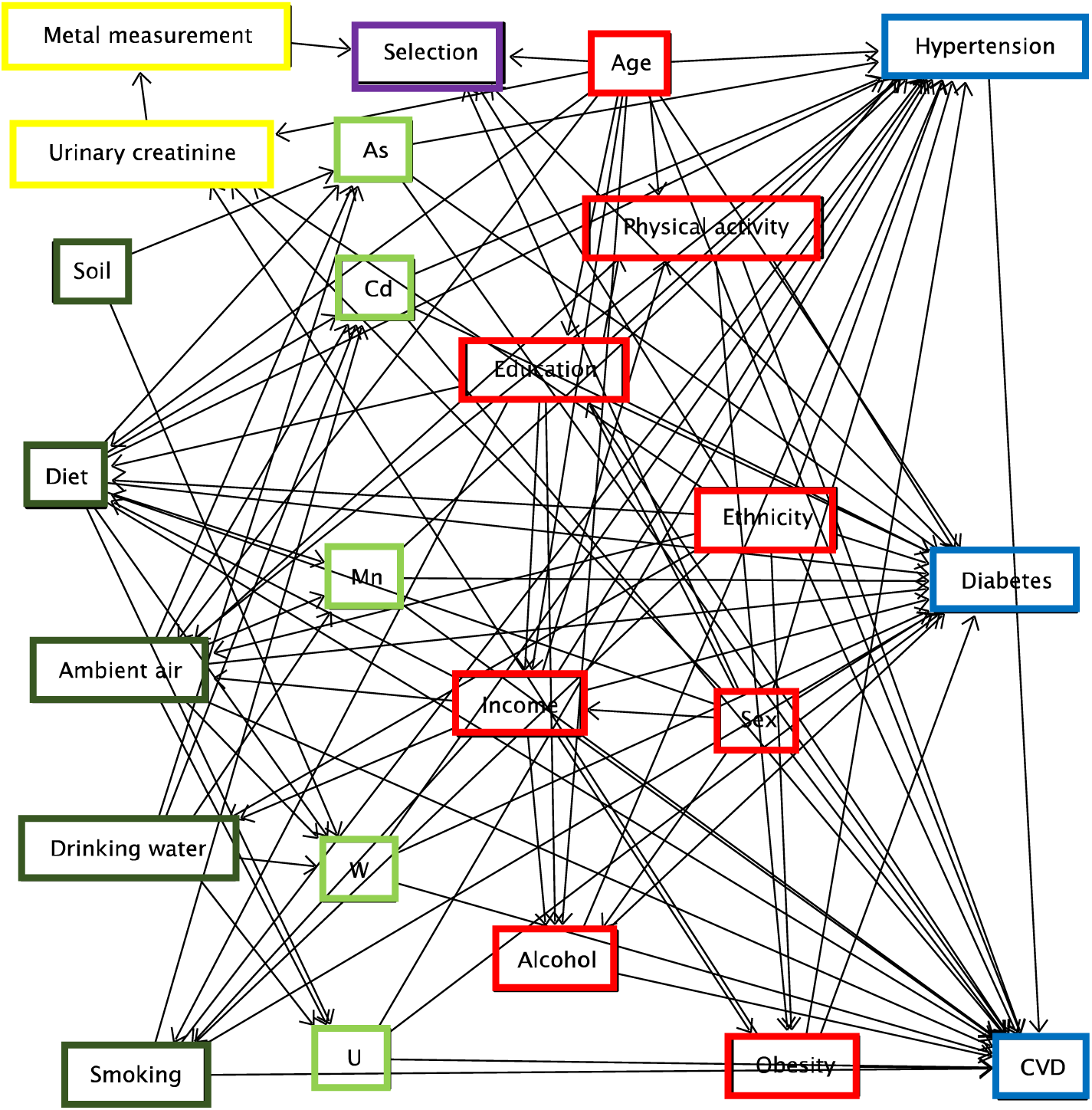
Directed acyclic graph representing the putative causal structure for the association between metal mixtures and cardiometabolic outcomes. Light green nodes represent exposures (i.e., metals), dark green nodes represent sources of exposure, blue nodes represent outcomes, red nodes represent risk factors for the outcomes, yellow nodes represent measurement-related nodes, and the purple node represents selection into the study. Figure created in DAGitty. CVD = cardiovascular disease; As = arsenic; Cd = cadmium; Mn = manganese; W = tungsten; U = uranium.

### Evaluation and application data

To evaluate and apply the evidence-based DAG, we used data from the San Luis Valley Diabetes Study (SLVDS), a prospective cohort study assessing the risk factors for chronic disease among Hispanic and non-Hispanic white adults in Alamosa or Conejos counties, Colorado. Data collection methods have been detailed elsewhere (Hamman et al. 1989). Briefly, the study population was recruited in two phases (Phase 1: 1984-1985; Phase 2: 1986-1987). People with diabetes were recruited through medical record reviews and local advertisements. People without diabetes were recruited using a stratified random sampling scheme based on residential location in these counties. All participants (with or without diabetes) met three additional eligibility criteria: (1) aged 20-74 years old, (2) able to provide informed consent, and (3) proficient in English or Spanish. Overall, 1823 individuals participated in the SLVDS, of whom 1795 had viable urine samples and were eligible for this analysis. Having a metal measurement was defined as having viable urine sample measured for metals (n = 1609).

Samples of urinary metals (approximately 120 mL) were collected and stored in trace metal free tubes in a freezer at −80°C until the laboratory analysis was conducted in 2008 and 2015 by the Colorado Department of Public Health and Environment chemistry laboratory. An inductively coupled argon plasma instrument with a mass spectrometer was used to detect the metal concentrations with a detection limit of 1 part in 10. Values below limit of detection were defined as the square root of detection limit divided by 2. All laboratory methods met the standards of the Clinical Laboratory Improvement Amendment and Environmental Protection Agency (Rivera-Núñez et al. 2012). Urinary creatinine (g/L) was quantified using a colorimetric assay by the Jaffe reaction (Delanghe and Speeckaert 2011).

The three outcome variables (i.e., cardiovascular disease, type 2 diabetes mellitus (hereafter ‘diabetes’), and hypertension) were assessed at baseline and dichotomized for this analysis. A participant was defined as having cardiovascular disease if they reported having had coronary bypass surgery or myocardial infarction. A participant was defined as having diabetes if they had a fasting venous plasma glucose ≥ 140 mg/dL (following a ≥8 hour fast), 2-hour venous plasma glucose ≥ 200 mg/dL (following consumption of a flavored drink), or were taking oral hypoglycemic medication (World Health Organization 1985). Measurements of fasting glucose used the glucose oxidase method with venous plasma (Hamman et al. 1989; Beckman Instruments 1988). A participant was defined as having hypertension if they had average systolic blood pressure ≥ 130 mmHg or average diastolic blood pressure ≥ 80 mmHg based on the average of the second and third blood pressure measurements (Whelton et al. 2018).

Other health and demographic data were collected at baseline. Researchers measured participants height and weight, and these measurements were used to calculate body mass index (BMI; obesity defined as BMI >30 kg/m^2^). Participants self-reported sex, age, educational attainment (<12 years/12 years/>12 years), ethnicity (Hispanic/non-Hispanic), annual gross household income (0-$14,999/$15,000-$34,999/≥$35,000), smoking status (never smoker [< 100 lifetime cigarettes]/former smoker [≥ 100 lifetime cigarettes but not currently smoking]/current smoker [≥ 100 lifetime cigarettes and currently smoking]), physical activity, and diet. Participants’ combined overall physical activity level accounting for activity during work (ranging from “sedentary” to “heavy physical work”) and non-work time (ranging from “practically none” to “great amount” of physical activity) was categorized as sedentary, somewhat active, moderately active, or most active (Swenson et al. 2005). Diet was based on 24-hour recall and food frequency questionnaire data (Marshall, Hamman, and Baxter 1991; Marshall, Weiss, and Hamman 1993). Micro and macronutrient intake was measured as g/day, and alcohol intake was measured as g/week. Since the macro and micronutrient variables (total kcals, vitamin C, zinc, selenium, vitamin A, beta carotene, folic acid, protein, total fats, saturated fats, monounsaturated fats, polyunsaturated fats, cholesterol, carbohydrates, total sugar, plant-based foods, insoluble, soluble, and total fiber, legumes, and omega-3) were highly correlated, we summarized them by conducting a principal component analysis. To do so, we first normalized the diet variables by centering and variance-standardizing them. Then, we used singular value decomposition implemented with the numpy linear algebra solver on the covariance of the normalized dietary data to identify the top two eigenvectors, which together accounted for 53% of the variance. We transformed the participants’ dietary data by matrix multiplication with the top two eigenvectors to project the dietary data onto the first two principal components.

### Statistical analysis

Our analysis was split into two parts: (1) evaluation of the DAG, and (2) application of the DAG in a pilot analysis. For both parts, we mapped each node in the evidence-based DAG to the corresponding variables in the SLVDS data. There were some exceptions; ambient air quality, drinking water quality, and soil exposure were unmeasured in the SLVDS. Selection was not evaluated in this analysis because selection into the study was an inclusion requirement.

In part one of our analysis (evaluation of the DAG), we first tested each conditional independence statement implied by the evidence-based DAG using linear (continuous outcomes), logistic (dichotomous outcomes), and ordered factor response logistic (ordinal outcomes). If the resulting *p* value for the coefficient from the model of the exposure and outcome conditional on the covariates was ≥ 0.05, we did not reject the null hypothesis that the conditional independence statement held. Because the hypothesis being tested is not that each individual conditional independence statement is true but that our DAG explains the associations observed in the data, we did not adjust for multiple testing. In fact, such adjustment would increase the *p* values and thus the fraction of conditional independence statements that are supported by the data. We assessed the proportion of the total testable conditional independence statements that were supported by the data. We then repeated this same process 100 more times, using DAGs with the same structure but randomly permuted nodes. If our DAG represents the real-world influences, then a lower proportion of the conditional independence statements generated by the DAGs with randomly permuted nodes should be supported by the data than the conditional independence statements generated by the evidence-based DAG. We assessed the fraction of the 100 DAGs with randomly permuted nodes for which this expectation held.

In part two of our analysis (pilot application of the DAG), we used DAGitty to identify the two minimally sufficient adjustment sets from the evidence-based DAG using the single door criterion (Pearl 2009). The exposure was combined As, Cd, Mn, W, and U. A minimal adjustment set was determined for each outcome separately. The sets were the same for all three outcomes and included: (1) ambient air, diet, drinking water, and smoking; and (2) ambient air, diet, ethnicity, income, and smoking (**Appendix B**). Minimal adjustment sets for each individual metal—outcome pair were also identified and reported in **Appendix B**. Since ambient air and drinking water exposures were not measured in this population, the sets used in the analysis were: (1) smoking and diet; and (2) ethnicity, income, smoking, and diet. We estimated BKMR models for the associations between urinary metals concentrations (natural log transformed total As, Cd, Mn, U, and W; comparing the 75^th^ to the 25^th^ percentile) and the likelihood of having each outcome (one outcome per model). Because we were interested in assessing applicability of adjustment sets, we conducted complete-case analyses based on variables included in the models. Additional information on the number of participants missing data is provided in **Appendix C.** To try to mitigate potential bias due to unmeasured variables, we conducted a sensitivity analysis additionally adjusting for other variables in the DAG (i.e., sex, age, obesity, and urinary creatinine). Analyses were conducted in R (R Core Team, Vienna, Austria) using the package BKMR (Bobb et al. 2015; 2018). Figures were developed using DAGitty and ggplot2 in R (Textor et al. 2016; Wickham 2009).

## Results

### DAG development

We identified 42 articles that met the criteria for inclusion in the literature review (**Figure 1**). These articles included six meta-analyses, 21 reviews, nine systematic reviews, and six combined systematic review and meta-analyses. The most commonly included metals were As (n = 22) and Cd (n = 17; **Table 1**). The evidence-based DAG illustrating the putative causal structure relating metal mixture exposures to cardiometabolic outcomes is presented in **Figure 2**. The DAG illustrating the putative causal structure relating individual metals and cardiometabolic outcomes excluding variables excluded from this analysis is presented in **Supplemental Figure 1**.

**Table 1.**
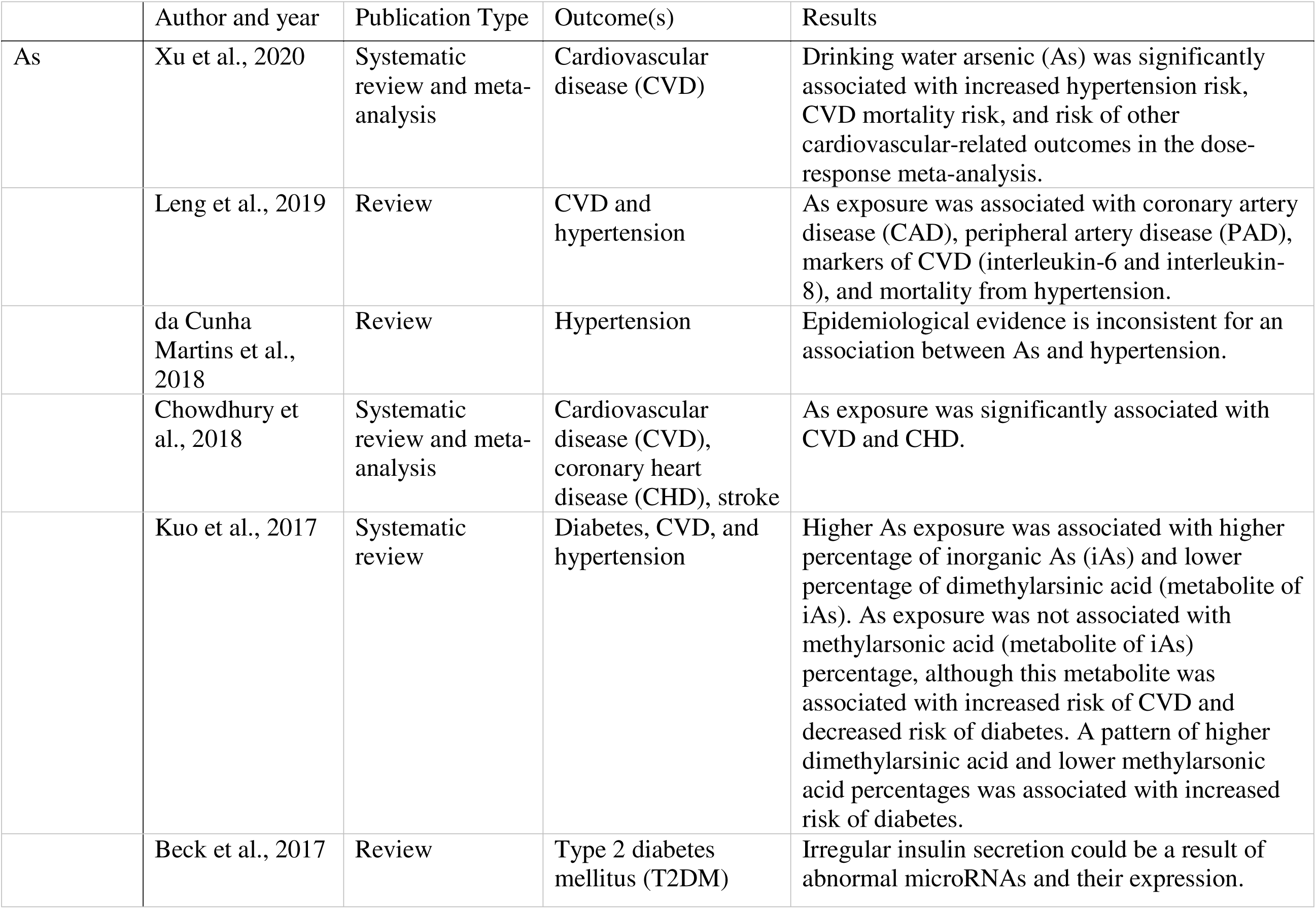
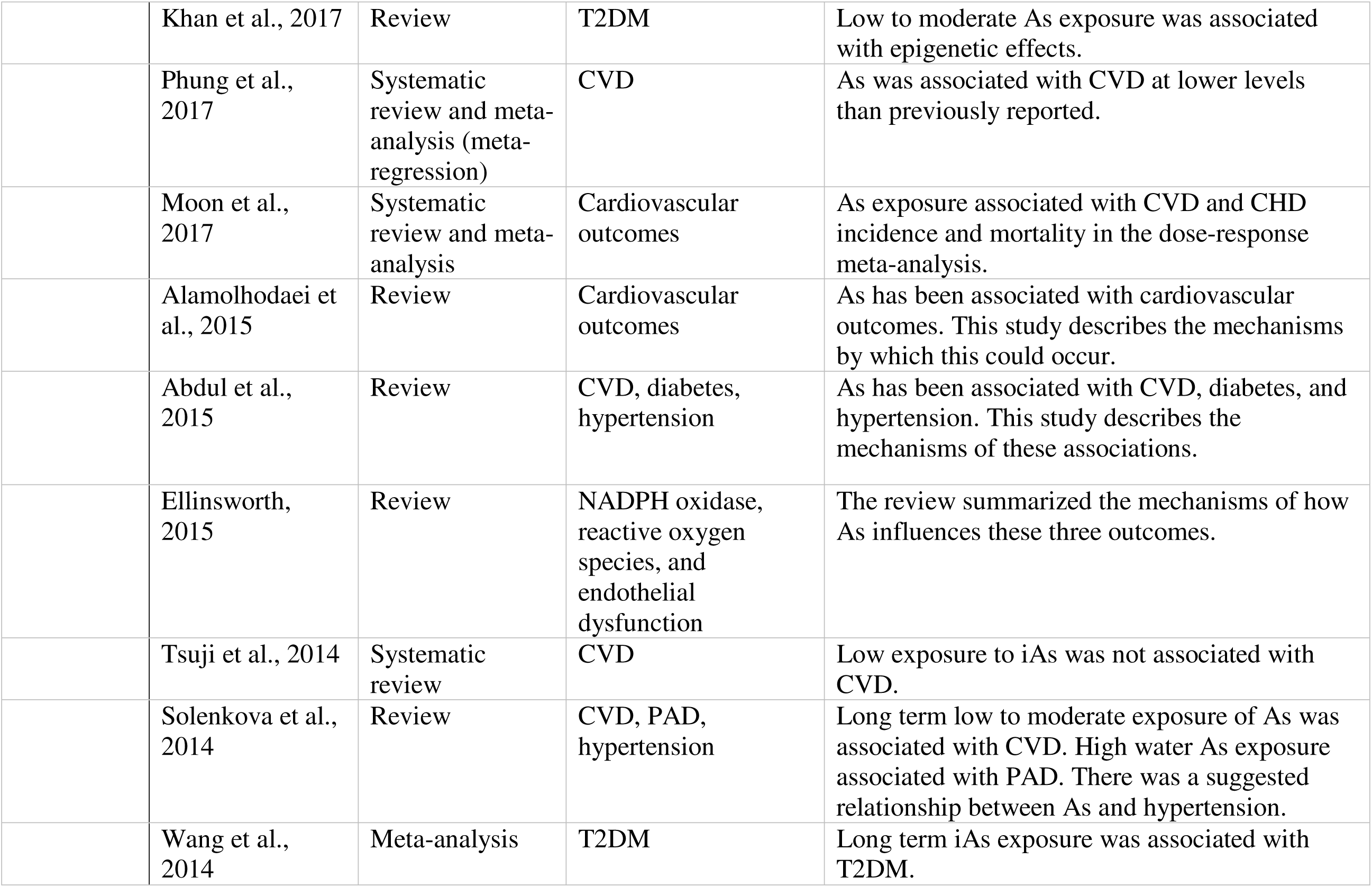
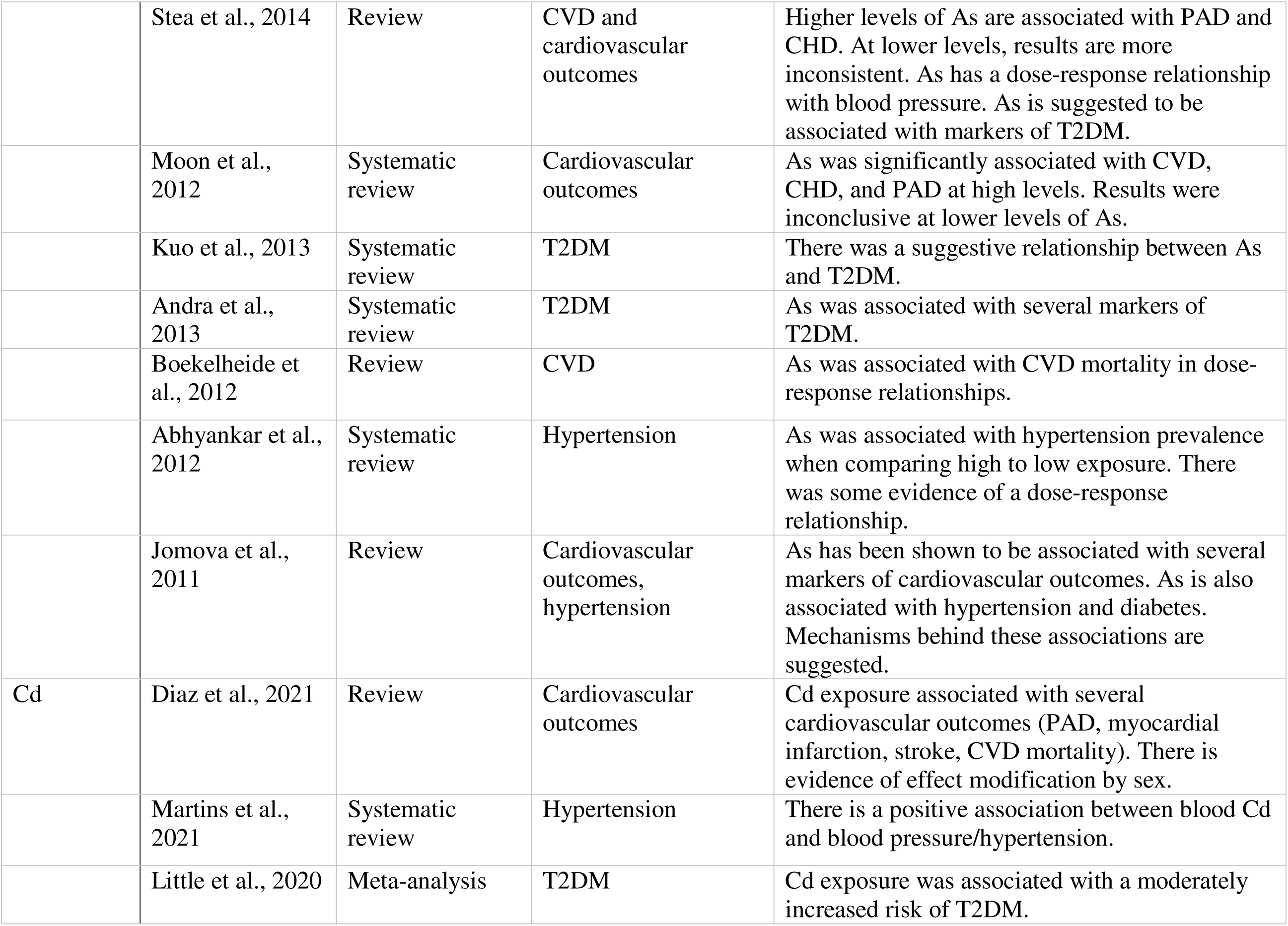
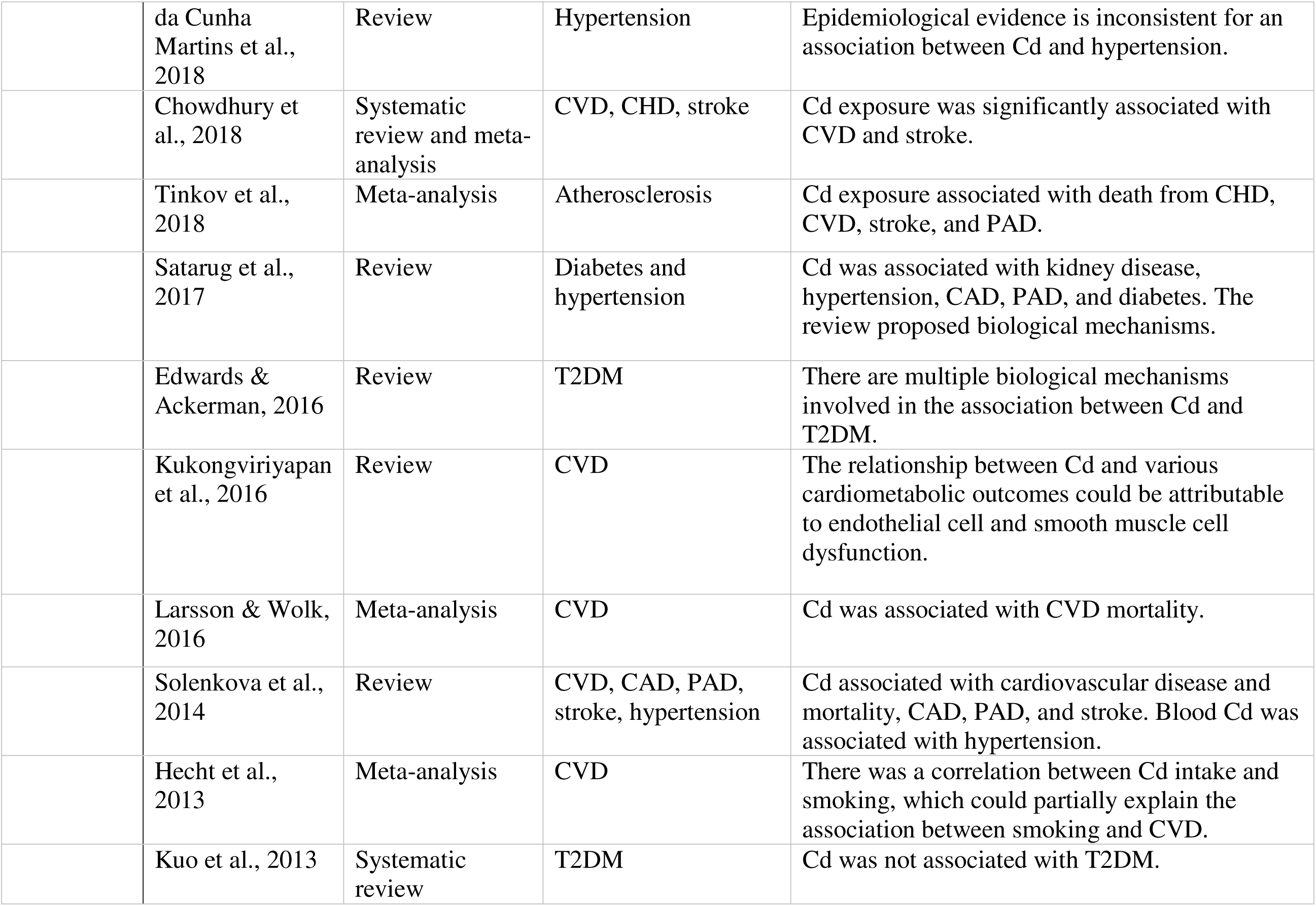
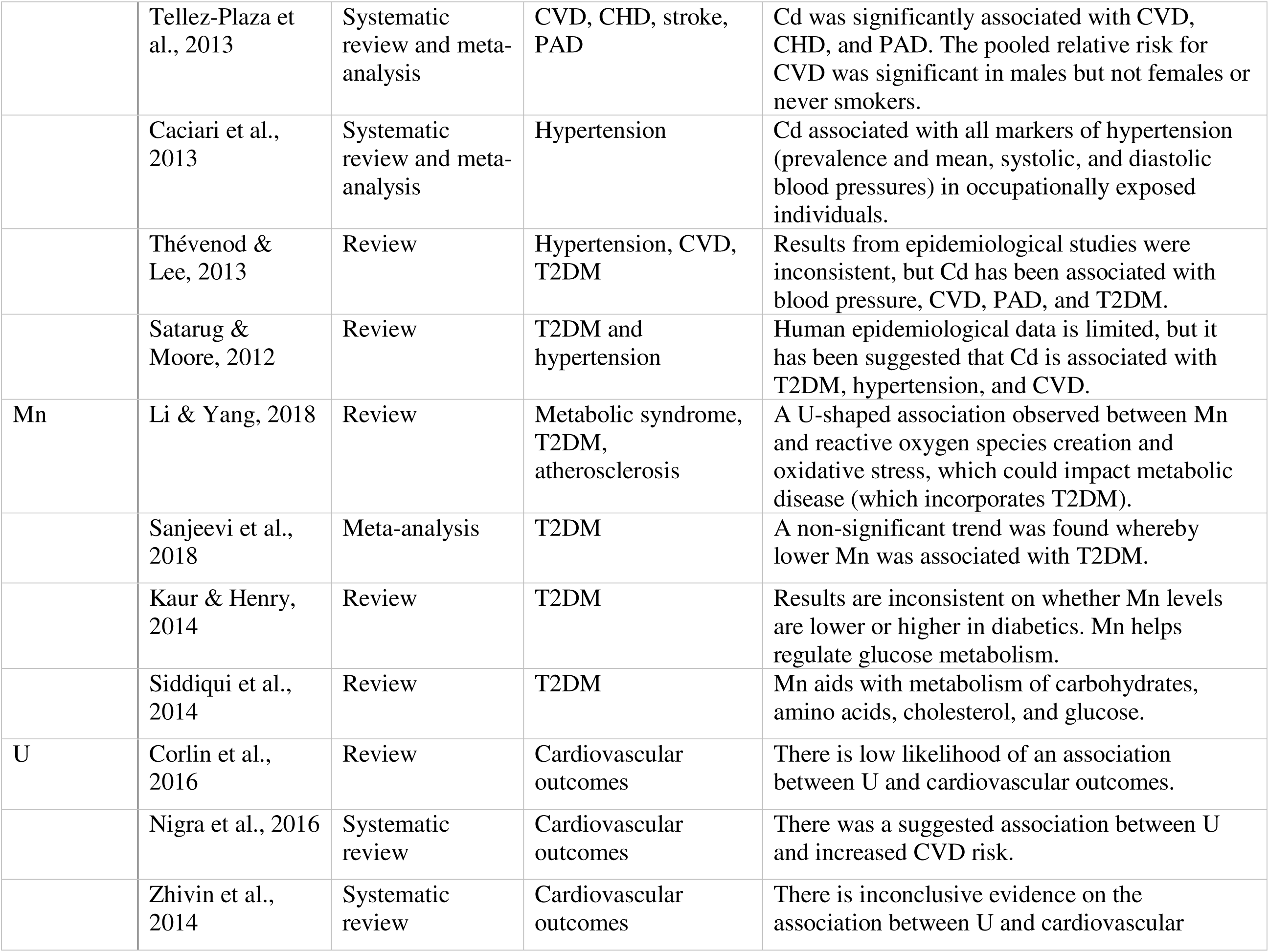
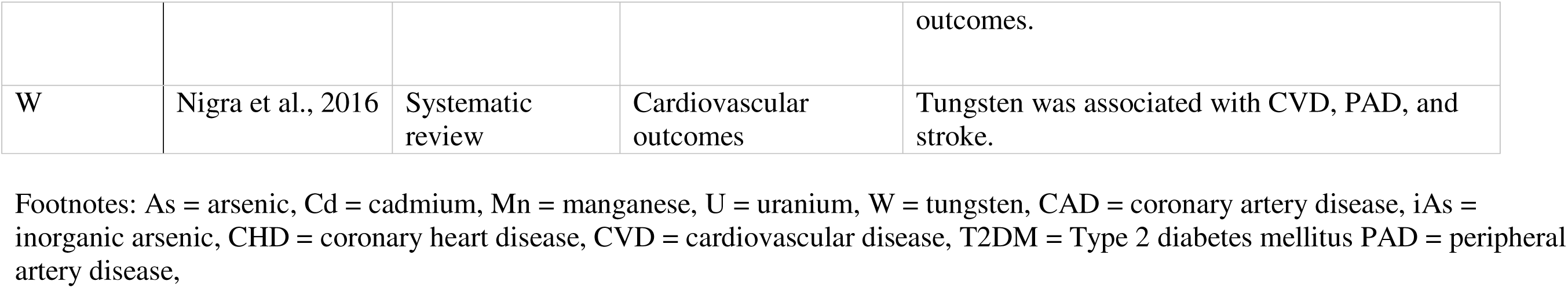
Literature review summary

|  | Author and year | Publication Type | Outcome(s) | Results |
| --- | --- | --- | --- | --- |
| As | Xu et al., 2020 | Systematic review and meta-analysis | Cardiovascular disease (CVD) | Drinking water arsenic (As) was significantly associated with increased hypertension risk, CVD mortality risk, and risk of other cardiovascular-related outcomes in the dose-response meta-analysis. |
|  | Leng et al., 2019 | Review | CVD and hypertension | As exposure was associated with coronary artery disease (CAD), peripheral artery disease (PAD), markers of CVD (interleukin-6 and interleukin-8), and mortality from hypertension. |
|  | da Cunha Martins et al., 2018 | Review | Hypertension | Epidemiological evidence is inconsistent for an association between As and hypertension. |
|  | Chowdhury et al., 2018 | Systematic review and meta-analysis | Cardiovascular disease (CVD), coronary heart disease (CHD), stroke | As exposure was significantly associated with CVD and CHD. |
|  | Kuo et al., 2017 | Systematic review | Diabetes, CVD, and hypertension | Higher As exposure was associated with higher percentage of inorganic As (iAs) and lower percentage of dimethylarsinic acid (metabolite of iAs). As exposure was not associated with methylarsonic acid (metabolite of iAs) percentage, although this metabolite was associated with increased risk of CVD and decreased risk of diabetes. A pattern of higher dimethylarsinic acid and lower methylarsonic acid percentages was associated with increased risk of diabetes. |
|  | Beck et al., 2017 | Review | Type 2 diabetes mellitus (T2DM) | Irregular insulin secretion could be a result of abnormal microRNAs and their expression. |
|  | Khan et al., 2017 | Review | T2DM | Low to moderate As exposure was associated with epigenetic effects. |
|  | Phung et al., 2017 | Systematic review and meta-analysis (meta-regression) | CVD | As was associated with CVD at lower levels than previously reported. |
|  | Moon et al., 2017 | Systematic review and meta-analysis | Cardiovascular outcomes | As exposure associated with CVD and CHD incidence and mortality in the dose-response meta-analysis. |
|  | Alamolhodaie et al., 2015 | Review | Cardiovascular outcomes | As has been associated with cardiovascular outcomes. This study describes the mechanisms by which this could occur. |
|  | Abdul et al., 2015 | Review | CVD, diabetes, hypertension | As has been associated with CVD, diabetes, and hypertension. This study describes the mechanisms of these associations. |
|  | Ellinsworth, 2015 | Review | NADPH oxidase, reactive oxygen species, and endothelial dysfunction | The review summarized the mechanisms of how As influences these three outcomes. |
|  | Tsuji et al., 2014 | Systematic review | CVD | Low exposure to iAs was not associated with CVD. |
|  | Solenkova et al., 2014 | Review | CVD, PAD, hypertension | Long term low to moderate exposure of As was associated with CVD. High water As exposure associated with PAD. There was a suggested relationship between As and hypertension. |
|  | Wang et al., 2014 | Meta-analysis | T2DM | Long term iAs exposure was associated with T2DM. |
|  | Stea et al., 2014 | Review | CVD and cardiovascular outcomes | Higher levels of As are associated with PAD and CHD. At lower levels, results are more inconsistent. As has a dose-response relationship with blood pressure. As is suggested to be associated with markers of T2DM. |
|  | Moon et al., 2012 | Systematic review | Cardiovascular outcomes | As was significantly associated with CVD, CHD, and PAD at high levels. Results were inconclusive at lower levels of As. |
|  | Kuo et al., 2013 | Systematic review | T2DM | There was a suggestive relationship between As and T2DM. |
|  | Andra et al., 2013 | Systematic review | T2DM | As was associated with several markers of T2DM. |
|  | Boekelheide et al., 2012 | Review | CVD | As was associated with CVD mortality in dose-response relationships. |
|  | Abhyankar et al., 2012 | Systematic review | Hypertension | As was associated with hypertension prevalence when comparing high to low exposure. There was some evidence of a dose-response relationship. |
|  | Jomova et al., 2011 | Review | Cardiovascular outcomes, hypertension | As has been shown to be associated with several markers of cardiovascular outcomes. As is also associated with hypertension and diabetes. Mechanisms behind these associations are suggested. |
| Cd | Diaz et al., 2021 | Review | Cardiovascular outcomes | Cd exposure associated with several cardiovascular outcomes (PAD, myocardial infarction, stroke, CVD mortality). There is evidence of effect modification by sex. |
|  | Martins et al., 2021 | Systematic review | Hypertension | There is a positive association between blood Cd and blood pressure/hypertension. |
|  | Little et al., 2020 | Meta-analysis | T2DM | Cd exposure was associated with a moderately increased risk of T2DM. |
|  | da Cunha Martins et al., 2018 | Review | Hypertension | Epidemiological evidence is inconsistent for an association between Cd and hypertension. |
|  | Chowdhury et al., 2018 | Systematic review and meta-analysis | CVD, CHD, stroke | Cd exposure was significantly associated with CVD and stroke. |
|  | Tinkov et al., 2018 | Meta-analysis | Atherosclerosis | Cd exposure associated with death from CHD, CVD, stroke, and PAD. |
|  | Satarug et al., 2017 | Review | Diabetes and hypertension | Cd was associated with kidney disease, hypertension, CAD, PAD, and diabetes. The review proposed biological mechanisms. |
|  | Edwards & Ackerman, 2016 | Review | T2DM | There are multiple biological mechanisms involved in the association between Cd and T2DM. |
|  | Kukongviriyapan et al., 2016 | Review | CVD | The relationship between Cd and various cardiometabolic outcomes could be attributable to endothelial cell and smooth muscle cell dysfunction. |
|  | Larsson & Wolk, 2016 | Meta-analysis | CVD | Cd was associated with CVD mortality. |
|  | Solenkova et al., 2014 | Review | CVD, CAD, PAD, stroke, hypertension | Cd associated with cardiovascular disease and mortality, CAD, PAD, and stroke. Blood Cd was associated with hypertension. |
|  | Hecht et al., 2013 | Meta-analysis | CVD | There was a correlation between Cd intake and smoking, which could partially explain the association between smoking and CVD. |
|  | Kuo et al., 2013 | Systematic review | T2DM | Cd was not associated with T2DM. |
|  | Tellez-Plaza et al., 2013 | Systematic review and meta-analysis | CVD, CHD, stroke, PAD | Cd was significantly associated with CVD, CHD, and PAD. The pooled relative risk for CVD was significant in males but not females or never smokers. |
|  | Caciari et al., 2013 | Systematic review and meta-analysis | Hypertension | Cd associated with all markers of hypertension (prevalence and mean, systolic, and diastolic blood pressures) in occupationally exposed individuals. |
|  | Thévenod & Lee, 2013 | Review | Hypertension, CVD, T2DM | Results from epidemiological studies were inconsistent, but Cd has been associated with blood pressure, CVD, PAD, and T2DM. |
|  | Satarug & Moore, 2012 | Review | T2DM and hypertension | Human epidemiological data is limited, but it has been suggested that Cd is associated with T2DM, hypertension, and CVD. |
| Mn | Li & Yang, 2018 | Review | Metabolic syndrome, T2DM, atherosclerosis | A U-shaped association observed between Mn and reactive oxygen species creation and oxidative stress, which could impact metabolic disease (which incorporates T2DM). |
|  | Sanjeevi et al., 2018 | Meta-analysis | T2DM | A non-significant trend was found whereby lower Mn was associated with T2DM. |
|  | Kaur & Henry, 2014 | Review | T2DM | Results are inconsistent on whether Mn levels are lower or higher in diabetics. Mn helps regulate glucose metabolism. |
|  | Siddiqui et al., 2014 | Review | T2DM | Mn aids with metabolism of carbohydrates, amino acids, cholesterol, and glucose. |
| U | Corlin et al., 2016 | Review | Cardiovascular outcomes | There is low likelihood of an association between U and cardiovascular outcomes. |
|  | Nigra et al., 2016 | Systematic review | Cardiovascular outcomes | There was a suggested association between U and increased CVD risk. |
|  | Zhivin et al., 2014 | Systematic review | Cardiovascular outcomes | There is inconclusive evidence on the association between U and cardiovascular outcomes. |
| W | Nigra et al., 2016 | Systematic review | Cardiovascular outcomes | Tungsten was associated with CVD, PAD, and stroke. |

Footnotes: As = arsenic, Cd = cadmium, Mn = manganese, U = uranium, W = tungsten, CAD = coronary artery disease, iAs = inorganic arsenic, CHD = coronary heart disease, CVD = cardiovascular disease, T2DM = Type 2 diabetes mellitus PAD = peripheral artery disease,

### SLVDS sample description

In the analytic sample, there were 1795 participants; however, the sample size for each analysis varied by the availability of data for the variables included in the specific model. Of all participants with metals measurements (n = 1609), 53% were female and 48% were Hispanic (**Table 2**). The mean age was 54 years (standard deviation = 12 years), and 58% had one of the three cardiometabolic outcomes at baseline. As shown in **Table 3**, the urinary metal concentrations in the SLVDS participants were higher than those reported in the 1988-1994 and 2015-2016 National Health and Nutrition Examination Surveys (NHANES) (“Fourth National Report on Human Exposure to Environmental Chemicals Update” 2019; Paschal et al. 1998).

**Table 2.** Characteristics of participants with metal exposure measurements at baseline

|  | n (%) or mean<br>(standard deviation) |
| --- | --- |
| Total | 1609 (100) |
| Sex |  |
| Female | 855 (53.1) |
| Male | 754 (46.9) |
| Age (years) | 54.3 (12.2) |
| Body mass index (kg/m<sup>2</sup>) | 26.7 (4.8) |
| Educational attainment |  |
| <12 years | 526 (32.7) |
| 12 years | 540 (33.6) |
| >12 years | 539 (33.5) |
| Ethnicity |  |
| Hispanic | 773 (48.0) |
| Non-Hispanic | 836 (52.0) |
| Income (\$) | |
| <15,000 | 664 (41.3) |
| 15,000 - <35,000 | 537 (33.4) |
| ≥35,000 | 287 (17.8) |
| Smoking status |  |
| Never | 721 (44.8) |
| Current | 387 (24.1) |
| Former | 499 (31.0) |
| Physical activity |  |
| Sedentary | 167 (10.4) |
| Somewhat active | 338 (21.0) |
| Moderately active | 457 (28.4) |
| Most active | 643 (40.0) |
| Alcohol intake (g/week) | 40.98 (104.1) |
| Caloric intake (kcal/day) | 1510.41 (577.9) |
| Outcome prevalence |  |
| Cardiovascular disease | 111 (6.9) |
| Diabetes | 409 (25.4) |
| Hypertension | 797 (49.5) |

**Table 3.** Baseline geometric mean metal concentration (µg/L) among SLVDS participants compared with National Health and Nutrition Examination Survey (NHANES) 1988-1994 and NHANES 2015-2016 participants

|  | SLVDS<br>(1984-1986) | NHANES III<br>(1988-1994) <sup>a</sup> | NHANES<br>(2015-2016) <sup>b</sup> |
| --- | --- | --- | --- |
| Total Arsenic <sup>c</sup> | 16.27 | - | 5.96 |
| Male | 18.87 | - | 6.54 |
| Female | 14.28 | - | 5.46 |
| Non-Hispanic | 17.84 | - | 5.17 |
| Hispanic | 14.73 | - | 6.38 |
| Cadmium | 0.69 | - | 0.13 |
| Male | 0.71 | - | 0.12 |
| Female | 0.67 | - | 0.14 |
| Non-Hispanic | 0.61 | - | 0.12 |
| Hispanic | 0.78 | - | 0.11 |
| Manganese | 0.67 | 0.53 | - |
| Male | 0.71 | - | - |
| Female | 0.65 | - | - |
| Non-Hispanic | 0.65 | - | - |
| Hispanic | 0.70 | - | - |
| Tungsten | 0.33 | 0.70 | 0.07 |
| Male | 0.36 | - | 0.08 |
| Female | 0.30 | - | 0.06 |
| Non-Hispanic | 0.41 | - | 0.06 |
| Hispanic | 0.26 | - | 0.08 |
| Uranium | 0.01 | - | 0.01 |
| Male | 0.02 | - | 0.01 |
| Female | 0.01 | - | 0.01 |
| Non-Hispanic | 0.01 | - | 0.01 |
| Hispanic | 0.01 | - | 0.01 |

<sup>a</sup> Data from Paschal et al. 1998
<sup>b</sup> Data from Fourth National Report on Human Exposure to Environmental Chemicals Update
<sup>c</sup> All metal measurements were conducted in urine. NHANES 2015-2016 did not report urinary measurements of manganese. NHANES III only reported urinary metal measurements of manganese and tungsten for the overall population.

### DAG assessment

Our evidence-based DAG was largely consistent with the SLVDS data. The evidence-based DAG indicated 449 conditional independence statements, of which 163 were testable based on our available data. Of these, 64% of these were supported by the SLVDS data (*p* ≥ 0.05). The sample sizes for these tests varied from 1461 to 1780 participants. Of the 30 conditional independence statements implying a single metal exposure and single outcome (e.g., diabetes is conditionally independent of Cd given ethnicity, income, smoking, and diet), 90% were supported by the data. To test whether support for 64% of conditional independencies represents an elevated number, we compared the result against 100 DAGs with randomly permutated nodes. Only five of the random DAGs had more than 64% of conditional independence statements supported by the data (**Supplemental Figure 2**).

Testing our DAG with the SLVDS data, clear patterns emerged regarding the dependencies of specific variables on other variables. Several nodes were frequently found to be conditionally dependent on other nodes identified in the evidence-based DAG. For example, statements including having a metal measurement (i.e., having viable urine) all had a *p* ≥ 0.05, indicating that the dependencies specified in the DAG were supported by the data. Similarly, age, as expected according to the DAG, was conditionally dependent on all related nodes. Every metal was found to be conditionally independent from urinary creatinine (*p* < 0.05), but urinary creatinine was consistently conditionally dependent on other nodes (e.g., alcohol intake and diet). A detailed list of all conditional independence statements and the corresponding *p* values can be found in **Appendix D**.

### DAG application – pilot analysis

Using either minimally sufficient adjustment set identified by the evidence-based DAG, we did not observe strong evidence of an association between metal mixture exposures and the likelihood of having any of the three adverse cardiometabolic outcomes (**Figures 3A-C; Supplemental Figures 3A-C**). Several metals exhibited U-shaped associations, such as Mn with diabetes and Cd with hypertension. There was no indication of interaction among any metals within the mixture for any of the outcomes (**Supplemental Figure 4A-C**). Associations between metals and cardiometabolic outcomes remained null when additionally adjusting for sex, age, obesity, and urinary creatinine (**Supplemental Figure 5A-C**).

**Figure 3A.**
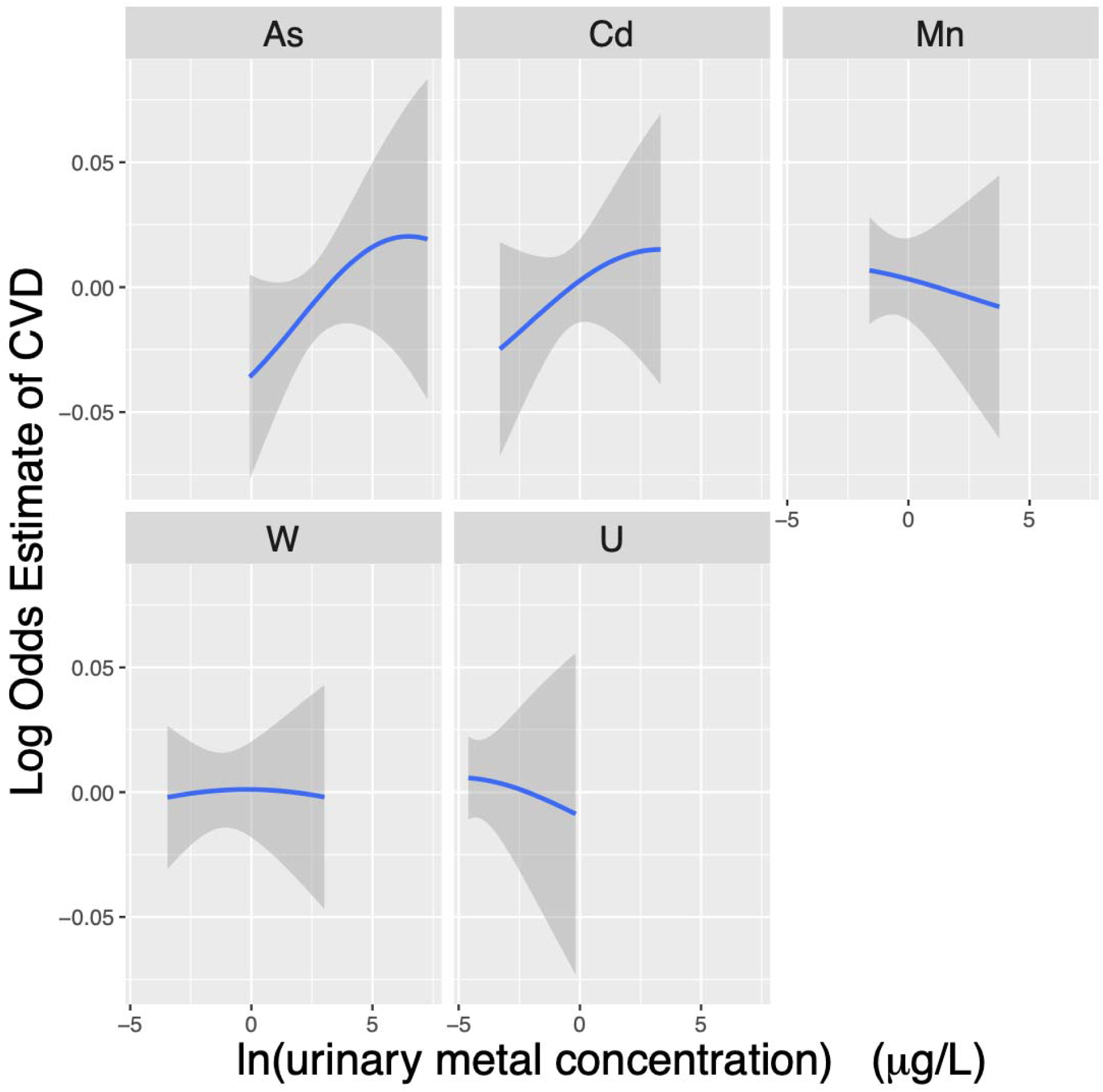
Exposure-response functions relating urinary metal concentrations to the log hazard estimate (interpreted as a log odds estimate) of prevalent cardiovascular disease (CVD). Model adjusted for ethnicity (Hispanic/non-Hispanic), income, smoking (never/former/current), and diet.

**Figure 3B.**
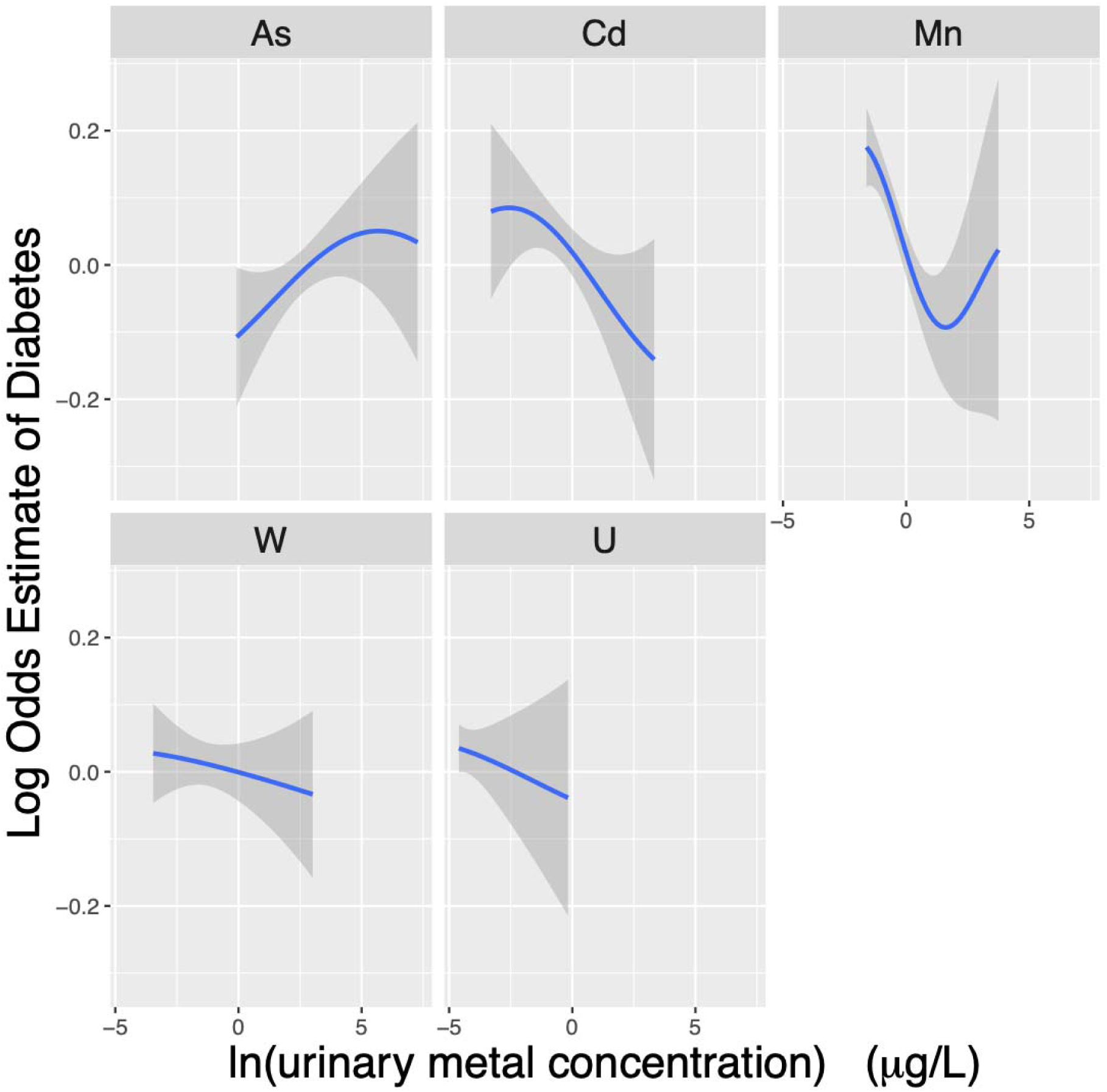
Exposure-response functions relating urinary metal concentrations to the log hazard estimate (interpreted as a log odds estimate) of prevalent diabetes. Model adjusted for ethnicity (Hispanic/non-Hispanic), income, smoking (never/former/current), and diet.

**Figure 3C.**
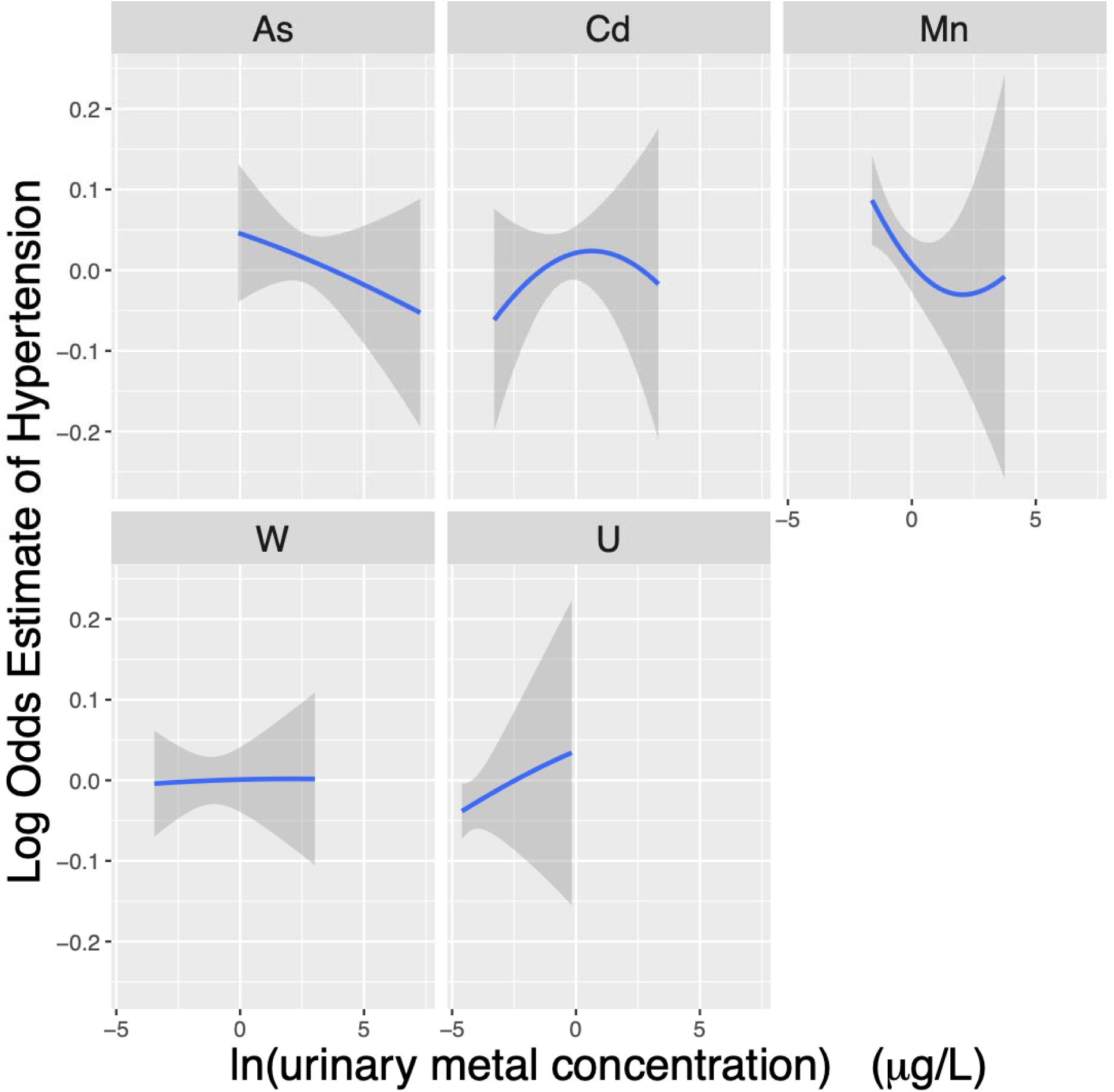
Exposure-response functions relating urinary metal concentrations to the log hazard estimate (interpreted as a log odds estimate) of prevalent hypertension. Model adjusted for ethnicity (Hispanic/non-Hispanic), income, smoking (never/former/current), and diet.

## Discussion and Conclusions

We developed, tested, and applied an evidence-based DAG illustrating the putative causal structure underlying associations between exposure to metal mixtures and cardiometabolic health. Our approach to developing the DAG using a systematic literature search was consistent with many of the tenants of DAG development discussed elsewhere (Corlin et al. 2018; Ferguson et al. 2020; Tennant et al. 2020). Our quantitative assessment of the DAG structure suggested that the evidence-based DAG was a reasonable representation of relationships among variables in a real data set. Furthermore, as demonstrated through our pilot analysis, environmental epidemiologists can apply our evidence-based DAG to transparently, reproducibly, and efficiently investigate associations among complex environmental exposures and cardiometabolic health outcomes using one of the proposed adjustment sets.

Through the literature review process, we identified several critical knowledge gaps that could be more effectively and efficiently addressed using our evidence-based DAG and analytic approach (e.g., assessing metal mixtures in relation to incident cardiometabolic health outcomes). Additional prospective studies of individual metals or metal mixtures and cardiometabolic outcomes are warranted, and evidence-based DAGs can guide the specification and interpretation of longitudinal models – even in complex metal mixture exposure scenarios (Corlin et al. 2016; Li and Yang 2018; Tyrrell et al. 2013). Insights into potential biological mechanisms can be derived from evidence-based DAGs, and these DAGs can be used to develop quantitative assessments of mechanistic hypotheses within observational studies (Corlin 2018). Additionally, further investigation into the mechanisms behind observed associations between metals and cardiometabolic outcomes could inform refinements of our evidence-based DAG (Beck, Styblo, and Sethupathy 2017; Edwards and Ackerman 2016; Khan et al. 2017). Future work could also investigate dose-response relationships between metal mixtures and health outcomes using the BKMR analytic approach we applied.

Beyond these challenges of multipollutant epidemiology that can be partially addressed through the development and application of evidence-based DAGs, we also identified several issues through our literature review that are unlikely to be handled by DAGs. DAGs can inform study design, exposure assessment priorities, and analytic model specification, but DAGs alone cannot fix fundamental problems with data collection (e.g., measurement error) or challenges of modeling environmental exposures over the life course. For example, several papers in our literature review discussed the use of urinary metal exposures as a limitation (notably one that is also present in our pilot analysis) because urinary exposures do not necessarily reflect total lifetime exposure, or even average exposure over an extended time period depending on the metal of interest and renal functioning (Balakrishnan et al. 2018; Larsson and Wolk 2016; Nong et al. 2016).

Our literature search suggested several mechanisms through which exposure to individual metals could potentially affect cardiometabolic health outcomes; however, future work is needed to understand how metals may interact physically and/or chemically to affect health. For individual metals, much of the literature focuses on how arsenic may be associated with cardiometabolic health through associations with modified gene expression, epigenetic changes, immune function, endothelial dysfunction, and oxidative stress (Andrew et al. 2008; Ellinsworth 2015; Khan et al. 2017). Similarly, there is extensive literature relating cadmium exposure to inflammatory biomarkers, kidney toxicity, endothelial dysfunction, and oxidative stress (Kukongviriyapan, Apaijit, and Kukongviriyapan 2016; Satarug, Vesey, and Gobe 2017). These types of hypotheses could be further explored using the evidence-based DAG we developed.

Applying the DAG we developed to epidemiological analyses will give environmental epidemiologists a defensible, reproducible method to identify and adjust for confounding. To the extent that new research is published challenging or adding to the DAG we show here, we encourage researchers to incorporate the new knowledge and update their adjustment sets; indeed, a prime advantage of using DAGs is that we can have a scientific conversation about explicit modeling assumptions (Tennant et al. 2020). For example, others may wish to include obesity as a cardiometabolic outcome (rather than a node that could be a potential confounder). Additionally, others may disagree with our inclusion (or exclusion) of certain studies supporting the presence or absence of specific arrows within the DAG (notably, we used evidence from meta-analyses, systematic reviews, and reviews rather than the primary literature). Nevertheless, given the similar results observed in our SLVDS analysis using each of the adjustment sets and given the DAG assessment results using the DAGs with randomly permutated nodes, we can suggest that our DAG development process was robust and that the DAG is compatible with real-world data. That said, we understand that a lack of evidence for an association does not guarantee a lack of true association (especially if the analyses were underpowered) – and this could have affected our assessment of the validity of the DAG. Other assumptions that we made while evaluating the DAG include that the shuffled DAGs (with randomly permuted nodes) were biologically plausible and that the conditional independence statements represent true relationships. The final DAG was highly complex due to the comprehensive search among exposures, outcomes, and other nodes. Future studies may consider assessing whether there is a simpler version of the DAG documenting relationships between metal mixtures and cardiometabolic outcomes, perhaps by determining how removing an edge or node affects other associations in the resulting structure. The aim of this study was to develop a DAG based on evidence from the literature and evaluate it using data; however, one could use data to develop a simpler DAG, although the results from data-driven methods must be evaluated within the context of current evidence.

The null results of the pilot BKMR analyses relating metal mixtures to cardiometabolic outcomes may be attributable to several factors. First, it is possible that the null trends overall reflect an averaging of positive and negative associations of components within the mixture. For example, As and Cd have been positively associated with diabetes, whereas Mn has been shown to aid in glucose metabolism and insulin secretion (Little et al. 2020; Siddiqui, Bawazeer, and Scaria Joy 2014; Wang et al. 2014). Similarly, results among studies in the literature review were conflicting for some metals; for example, the association between As and measures of cardiovascular disease was inconsistent across studies, particularly for lower exposure levels (K. A. Moon et al. 2017; Solenkova et al. 2014; Stea et al. 2014; Tsuji et al. 2014). We opted to include an arrow between a metal and outcome if at least one study from the review suggested an association; however, inconsistencies in previous literature highlight the need for additional research and can potentially explain the observed overall null association. Second, we used total arsenic rather than speciated arsenic. Certain species of inorganic arsenic have been found to be more toxic than forms of inorganic and organic arsenic (Ellinsworth 2015). Third, we did not have data on several nodes for the DAG (i.e., drinking water, soil, and ambient air quality). Exposure through ambient air is less common in rural areas with reduced industrial activity, such as the San Luis Valley where our participants resided (Briffa, Sinagra, and Blundell 2020). However, drinking water and soil are still potential sources of exposure in this population and should be investigated further. Fourth, the DAG assessment relied on the significance cutoff of 0.05 and did not consider effect sizes. Due to the varying sample sizes within the tests of conditional independence, we relied exclusively on *p* values rather than examining the magnitudes of effects. Fifth, the DAG assessment, while affirming that many relationships in the theoretical DAG were supported by the data, also revealed some conditionally dependent relationships that were not supported by the literature search. These associations (indicated by a *p* value less than 0.05) may warrant future examination; however, it is important to first identify mechanistic or theoretical support based in the literature before statistical investigation, as was done in the present study. Additionally, there are several mechanisms through which individual metals can be associated with certain cardiometabolic outcomes, and these could not be assessed in this pilot analysis. Furthermore, we note limitations of our pilot analysis such as high urinary metal concentrations in the study population compared to the U.S. population, the lack of diversity in our cohort beyond non-Hispanic and Hispanic white individuals, and the limited transportability of our results to non-rural populations. Although the pilot analysis results may not be transportable outside of populations like our target population, the results from our extensive literature review and evidence-based DAG could be applicable to any cohort.

Through our systematic literature search, we were able to construct an evidence-based DAG to inform adjustment set selection and/or study design for future longitudinal analyses of metal mixtures and cardiometabolic outcomes. We also demonstrated an approach to explicitly state and test assumptions through our application of the evidence-based DAG to the rich SLVDS data set. We encourage other environmental epidemiologists to develop and use such tools to increase the scientific rigor, transparency, and reproducibility of their work.

## Data Availability

The San Luis Valley Diabetes Study (SLVDS) summary data can be shared with the scientific community in accordance with the SLVDS data sharing plan (which includes Data Use Agreements and anonymizing of data to protect subject confidentiality). Analytic code is available upon reasonable request from the corresponding author.

## Abbreviations

As: arsenic
BKMR: Bayesian kernel machine regression
BMI: body mass index
CAD: coronary artery disease
Cd: cadmium
CHD: coronary heart disease
CVD: cardiovascular disease
DAG: directed acyclic graph
iAs: inorganic arsenic
Mn: manganese
NHANES: National Health and Nutrition Examination Surveys
PAD: peripheral artery disease
SLVDS: San Luis Valley Diabetes Study
T2DM: type 2 diabetes mellitus
U: uranium
W: tungsten

## Funding

Eunice Kennedy Shriver National Institute of Child Health & Human Development (NICHD) grant number K12HD092535 (Corlin), Tufts University Department of Public Health and Community Medicine (Corlin and Riseberg), Tufts Institute of the Environment (Riseberg), R00ES027853 (Alderete)

All the intellectual property and data generated were administered according to policies from the University of Colorado and the NIH, including the NIH Data Sharing Policy and Implementation Guidance of March 5, 2003. The San Luis Valley Diabetes Study (SLVDS) summary data can be shared with the scientific community in accordance with the SLVDS data sharing plan (which includes Data Use Agreements and anonymizing of data to protect subject confidentiality). Analytic code is available upon reasonable request from the corresponding author.

## Acknowledgements

We would like to thank the participants and staff of the San Luis Valley Diabetes Study.

## Notes

Author disclosures: The authors report no conflicts of interest.

### Competing Interest Statement

The authors have declared no competing interest.

### Author Declarations

All the intellectual property and data generated were administered according to policies from the University of Colorado and the NIH, including the NIH Data Sharing Policy and Implementation Guidance of March 5, 2003.

### Summary of Updates

We have modified the search methods and discussion.

