## Supplement for "Development and application of an evidence-based directed acyclic graph to evaluate the associations between metal mixtures and cardiometabolic outcomes"

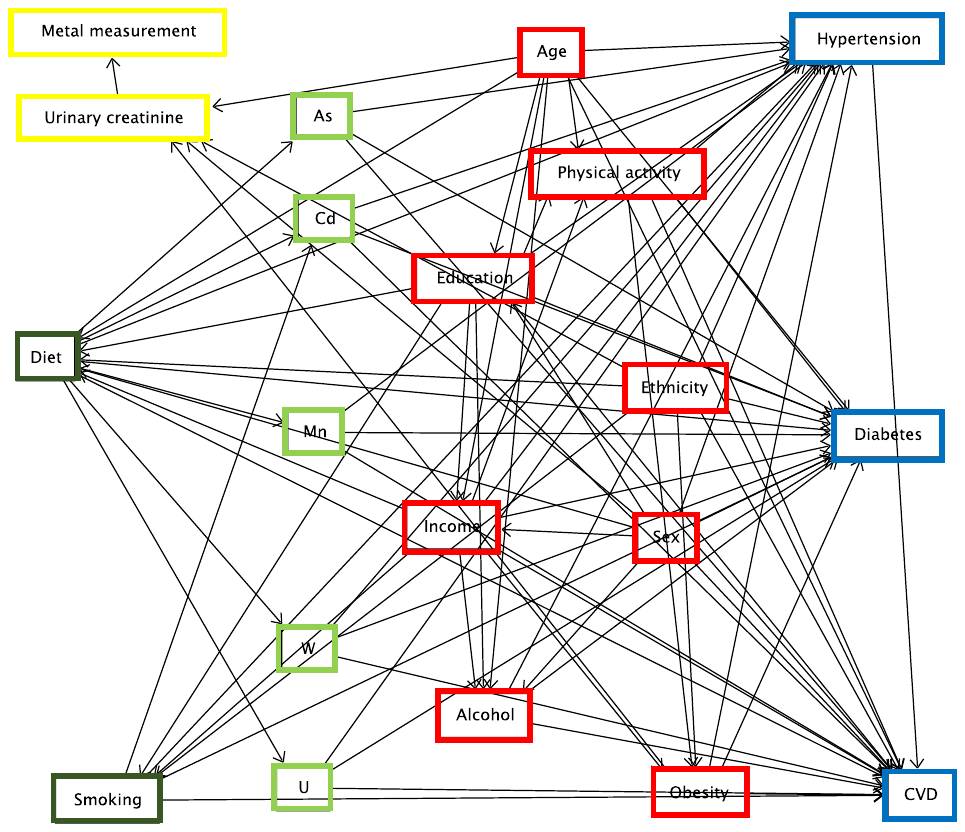

**Supplemental Figure 1**. Directed acyclic graph illustrating putative causal relationships among metals and cardiometabolic outcomes excluding variables excluded from the analysis

Footnotes: Light green nodes represent exposures (i.e., metals), dark green nodes represent sources of exposure, blue nodes represent outcomes, red nodes represent risk factors for the outcomes, and yellow nodes represent measurement-related nodes. Figure created in DAGitty. CVD = cardiovascular disease; As = arsenic; Cd = cadmium; Mn = manganese; W = tungsten; U = uranium.

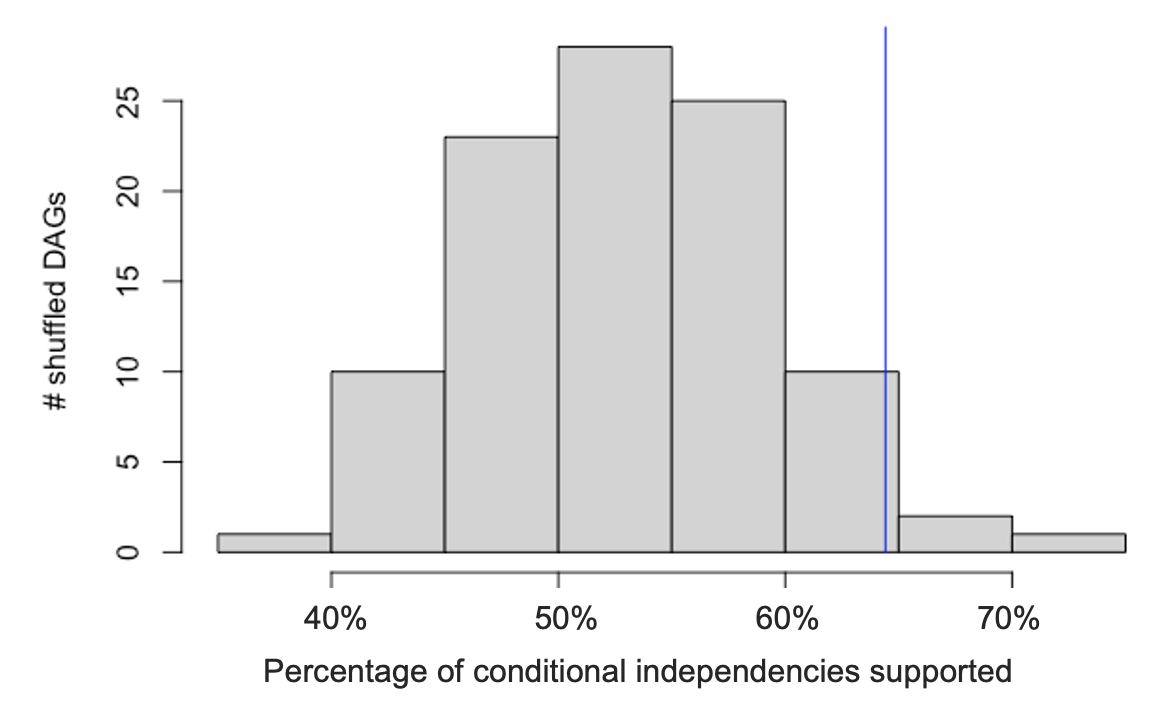

**Supplemental Figure 2**. Comparison of the number of conditional independence statements supported by the evidence-based DAG (64% of 163 testable statements; blue vertical line) versus the 100 DAGs with randomly permuted nodes (“shuffled DAGs”)

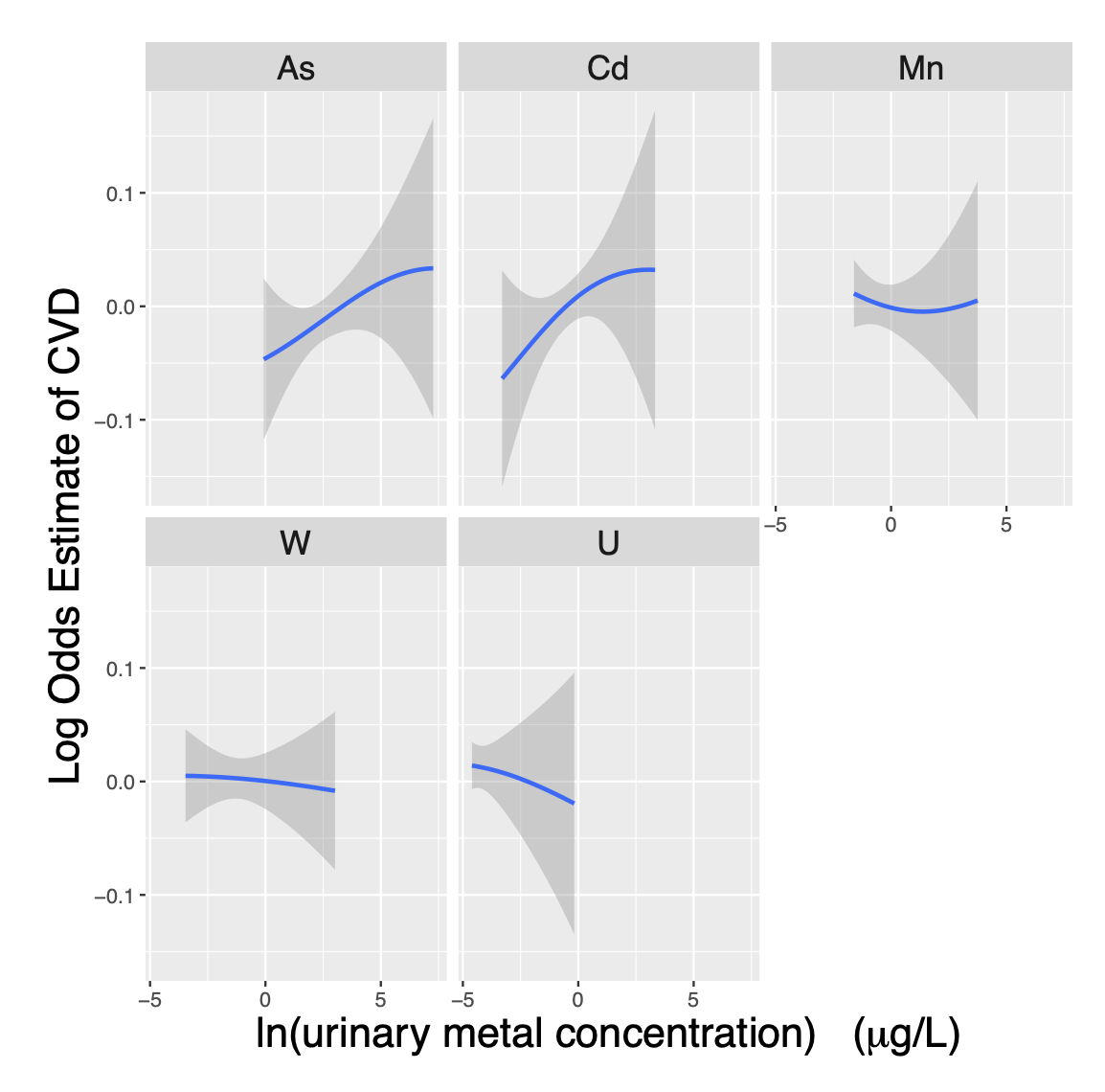

**Supplemental Figure 3A.** Exposure-response functions relating urinary metal concentrations to the log hazard estimate (interpreted as a log odds estimate) of prevalent cardiovascular disease (CVD). The model was adjusted for smoking (never/former/current) and diet.

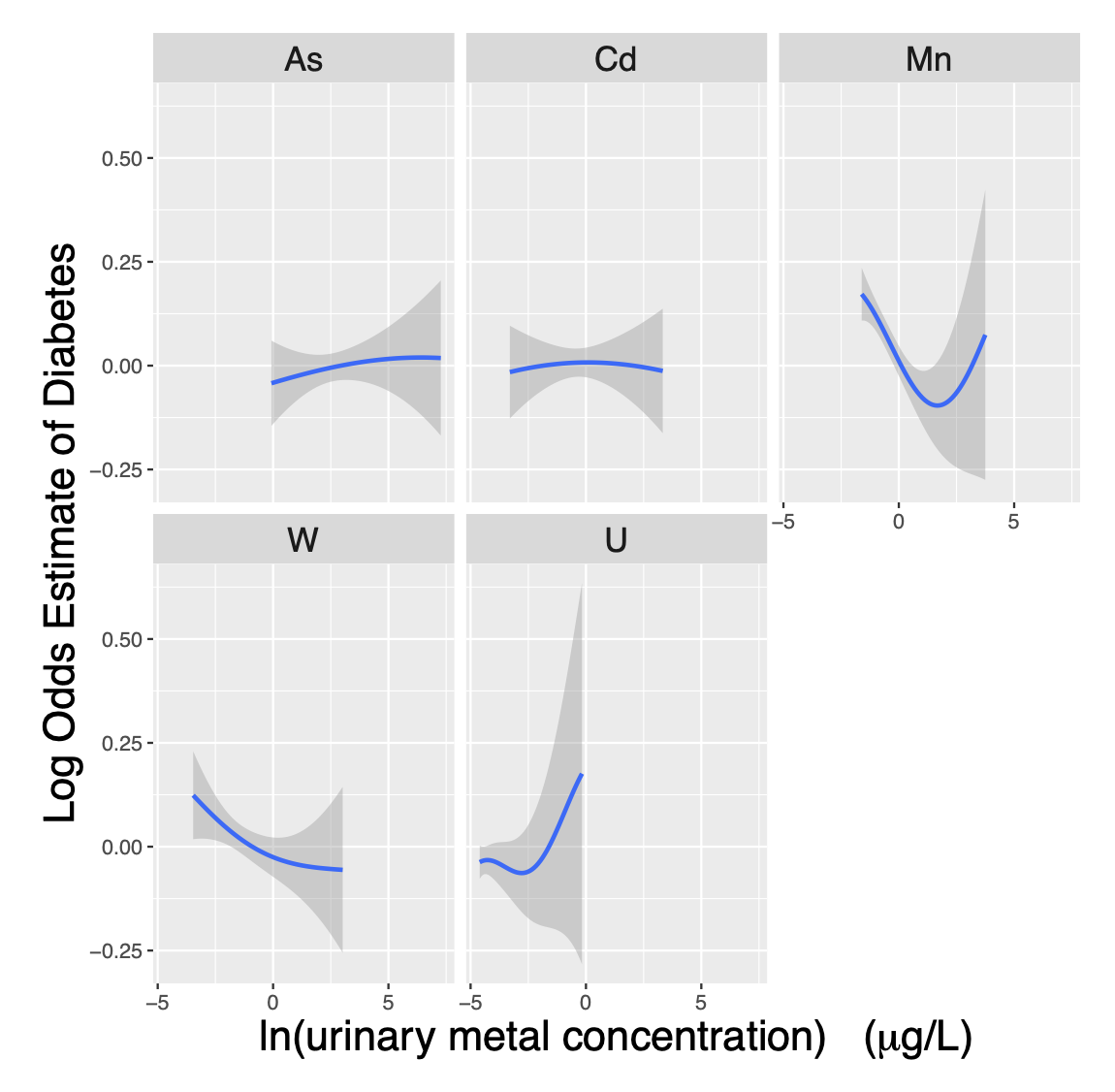

**Supplemental Figure 3B.** Exposure-response functions relating urinary metal concentrations to the log hazard estimate (interpreted as a log odds estimate) of prevalent diabetes. The model was adjusted for smoking (never/former/current) and diet.

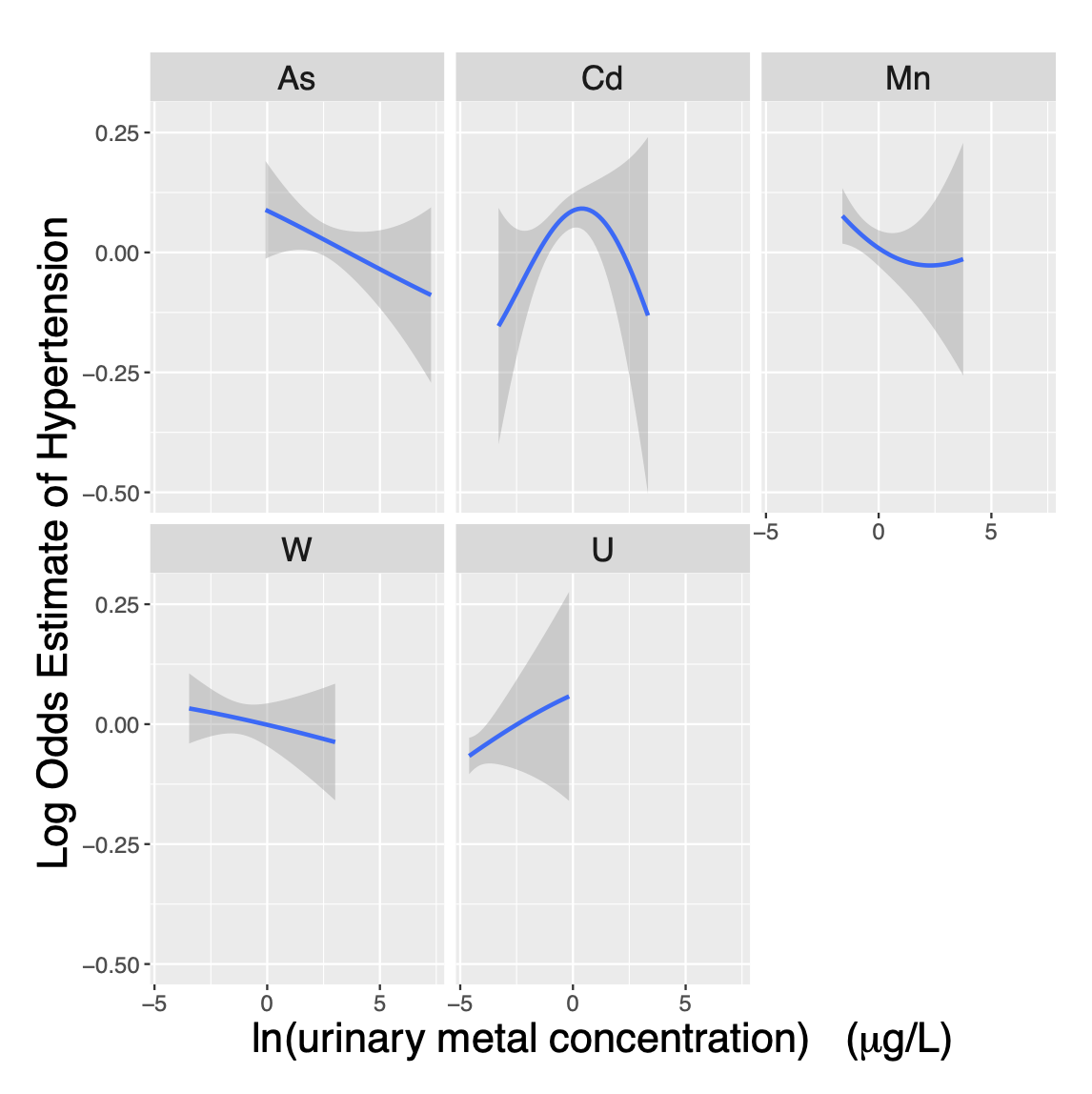

**Supplemental Figure 3C.** Exposure-response functions relating urinary metal concentrations to the log hazard estimate (interpreted as a log odds estimate) of prevalent hypertension. The model was adjusted for smoking (never/former/current) and diet.

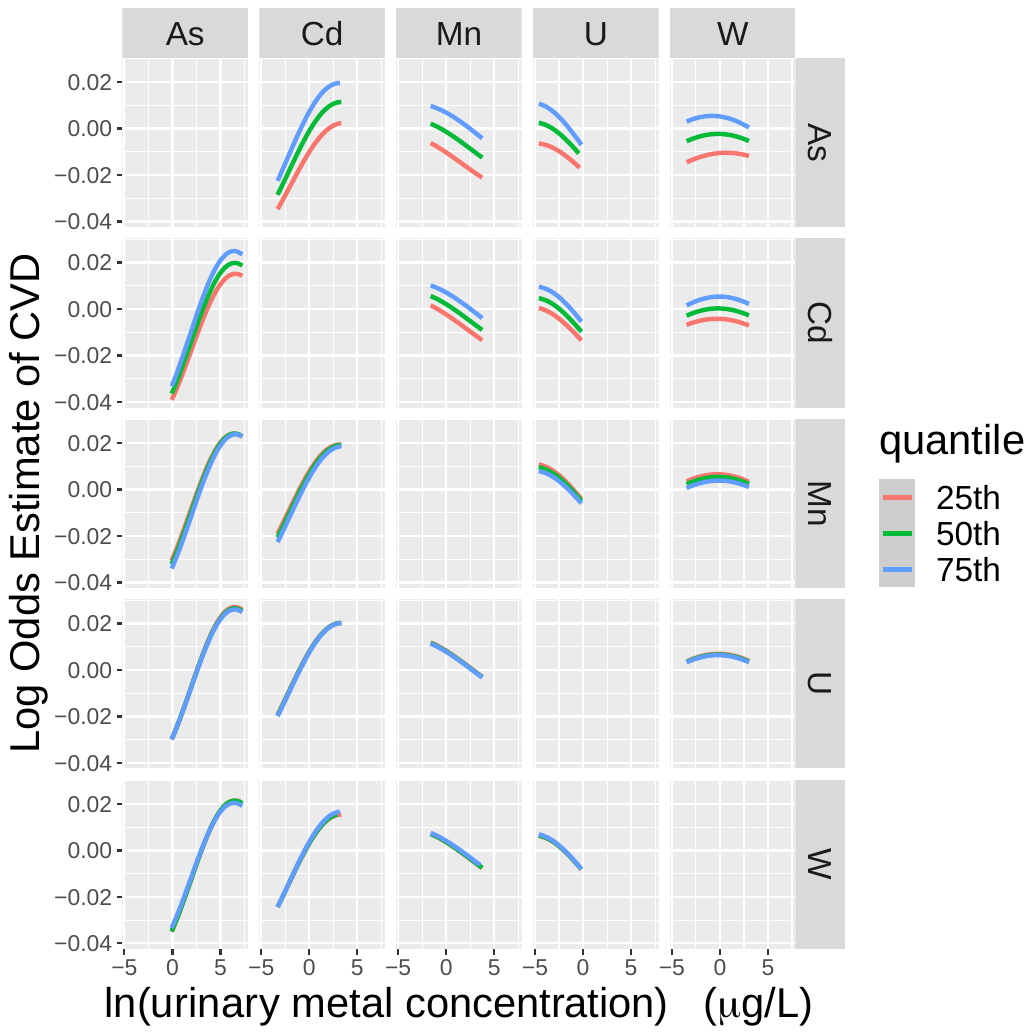

**Supplemental Figure 4A**. Exposure-response functions for the log hazard estimate (interpreted as a log odds estimate) of prevalent cardiovascular disease (CVD) for each metal (columns) when another metal (rows) is held constant at given quantiles and all other metals are held constant at their median. The model was adjusted for ethnicity (Hispanic/non-Hispanic), income, smoking (never/former/current), and diet.

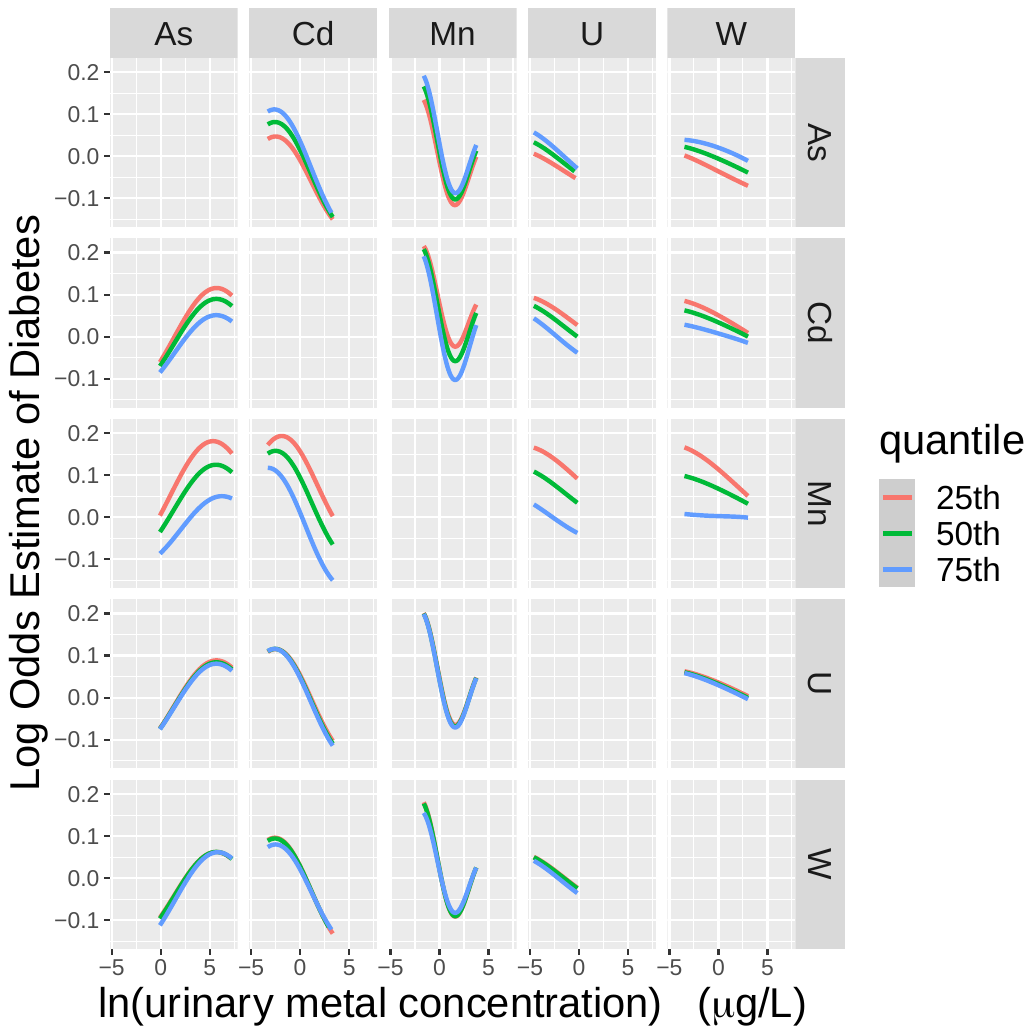

**Supplemental Figure 4B**. Exposure-response functions for the log hazard estimate (interpreted as a log odds estimate) of prevalent diabetes for each metal (columns) when another metal (rows) is held constant at given quantiles and all other metals are held constant at their median. The model was adjusted for ethnicity (Hispanic/non-Hispanic), income, smoking (never/former/current), and diet.

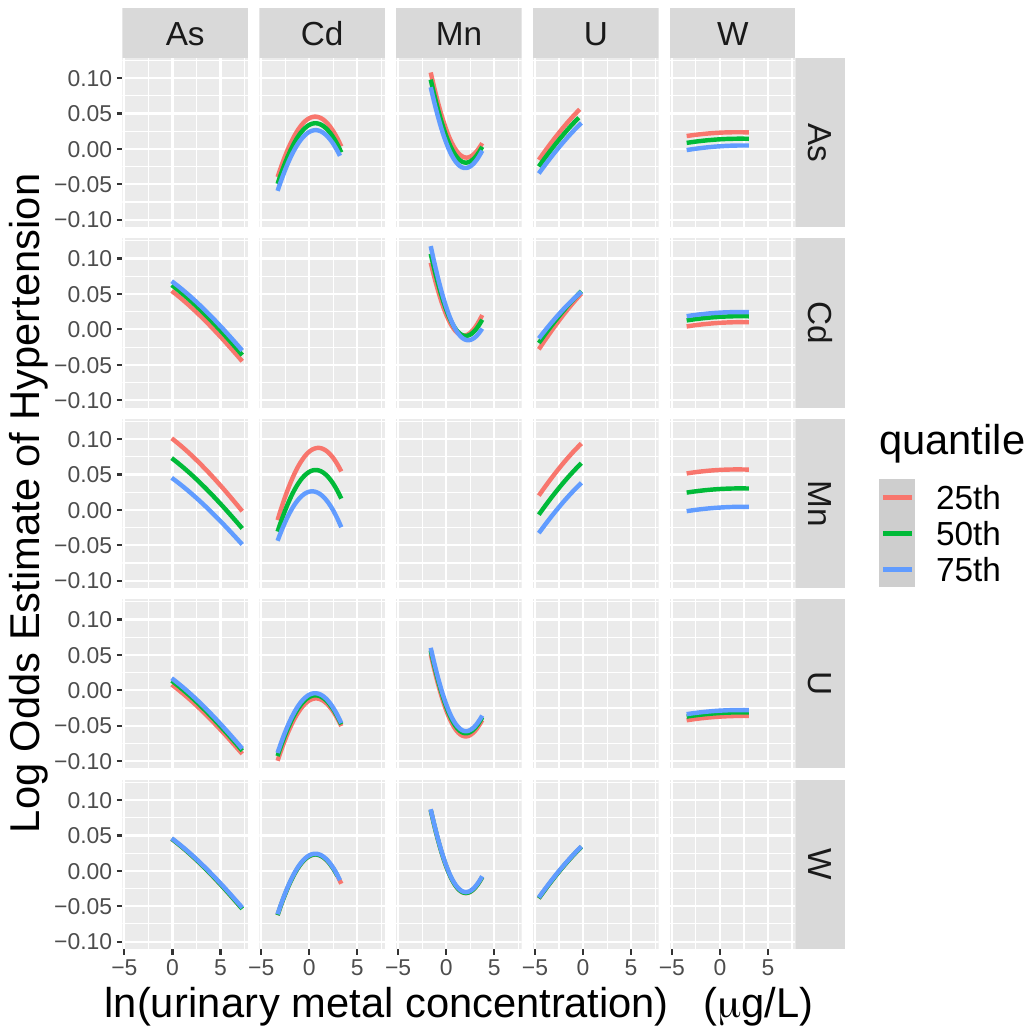

**Supplemental Figure 4C**. Exposure-response functions for the log hazard estimate (interpreted as a log odds estimate) of prevalent hypertension for each metal (columns) when another metal (rows) is held constant at given quantiles and all other metals are held constant at their median. The model was adjusted for ethnicity (Hispanic/non-Hispanic), income, smoking (never/former/current), and diet.

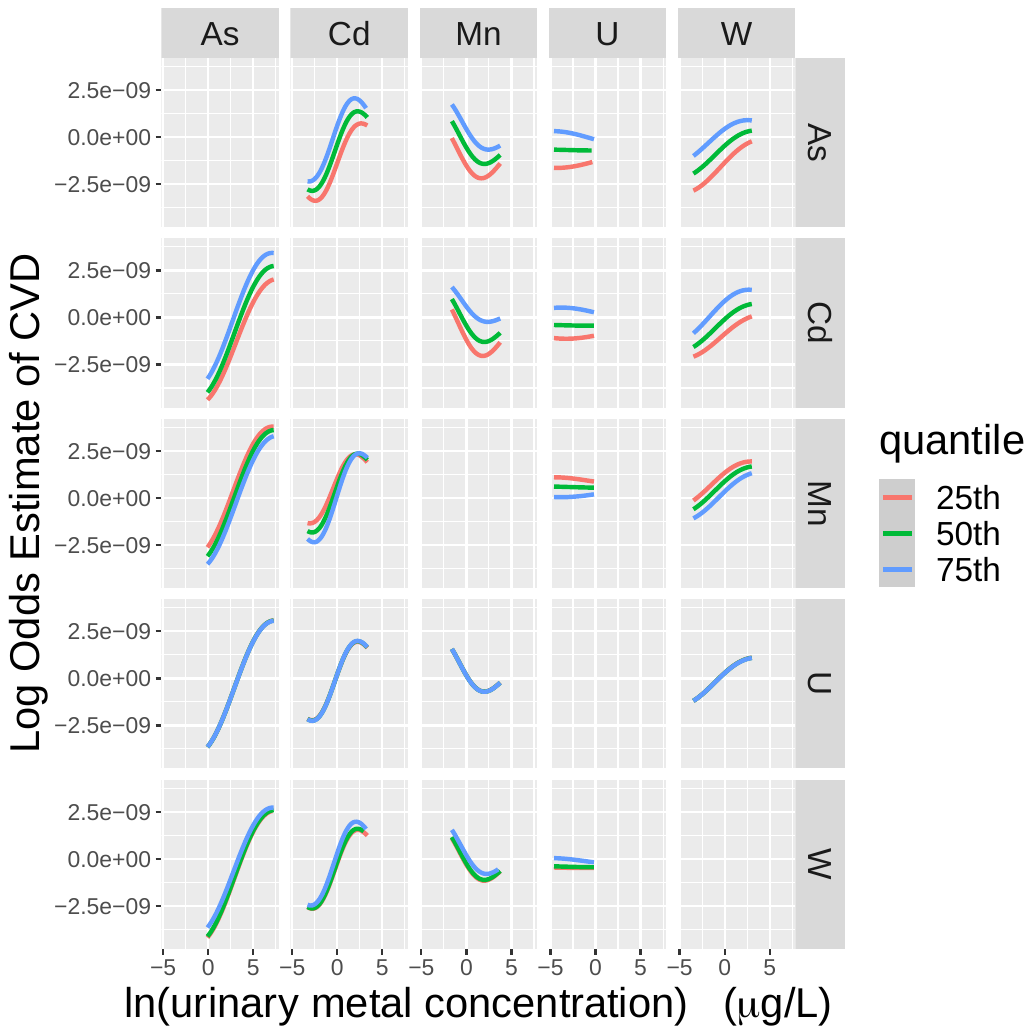

**Supplemental Figure 5A.** Exposure-response functions for the log hazard estimate (interpreted as a log odds estimate) of prevalent cardiovascular disease (CVD) for each metal (columns) when another metal (rows) is held constant at given quantiles and all other metals are held constant at their median. The model was adjusted for sex, age, ethnicity (Hispanic/non-Hispanic), income, obesity (BMI > 30 kg/m^2^), smoking (never/former/current), diet, and urinary creatinine (g/L).

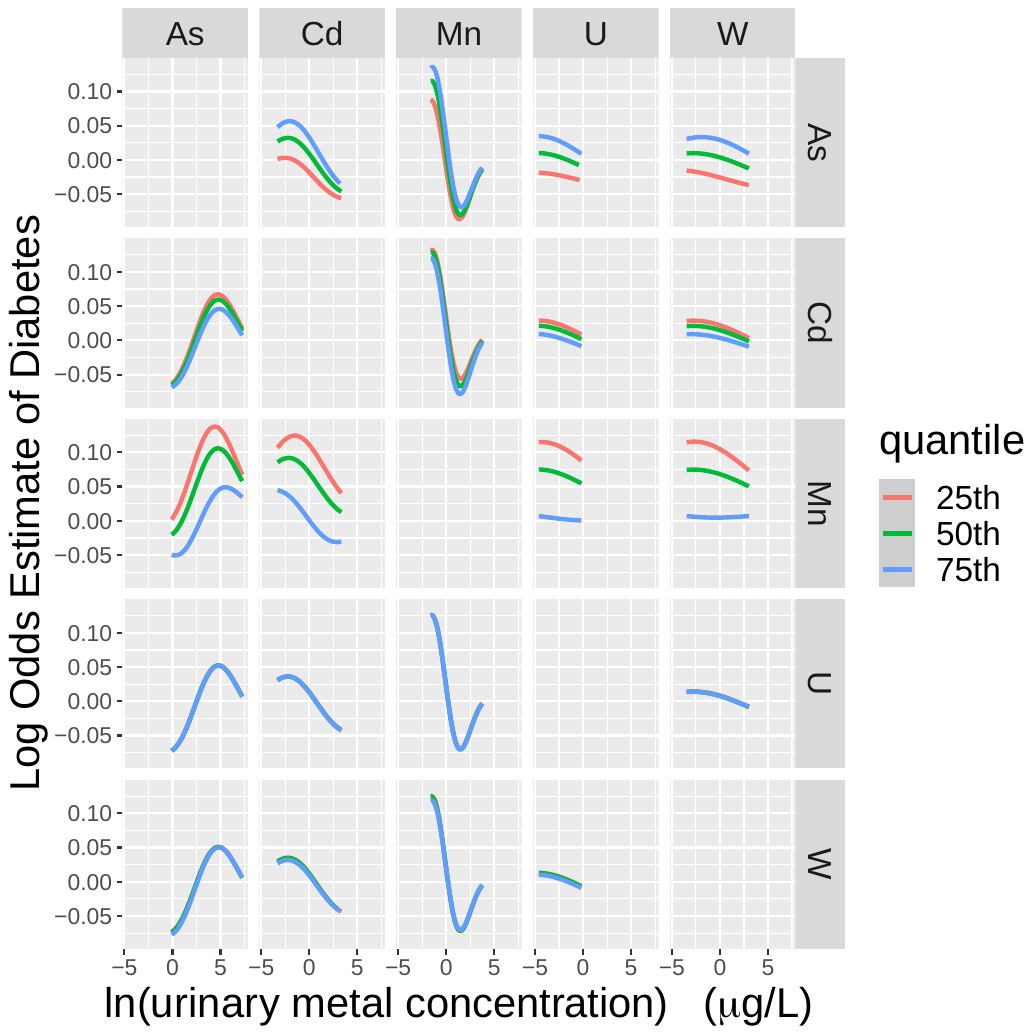

**Supplemental Figure 5B.** Exposure-response functions for the log hazard estimate (interpreted as a log odds estimate) of prevalent diabetes for each metal (columns) when another metal (rows) is held constant at given quantiles and all other metals are held constant at their median. The model was adjusted for sex, age, ethnicity (Hispanic/non-Hispanic), income, obesity (BMI > 30 kg/m^2^), smoking (never/former/current), diet, and urinary creatinine (g/L).

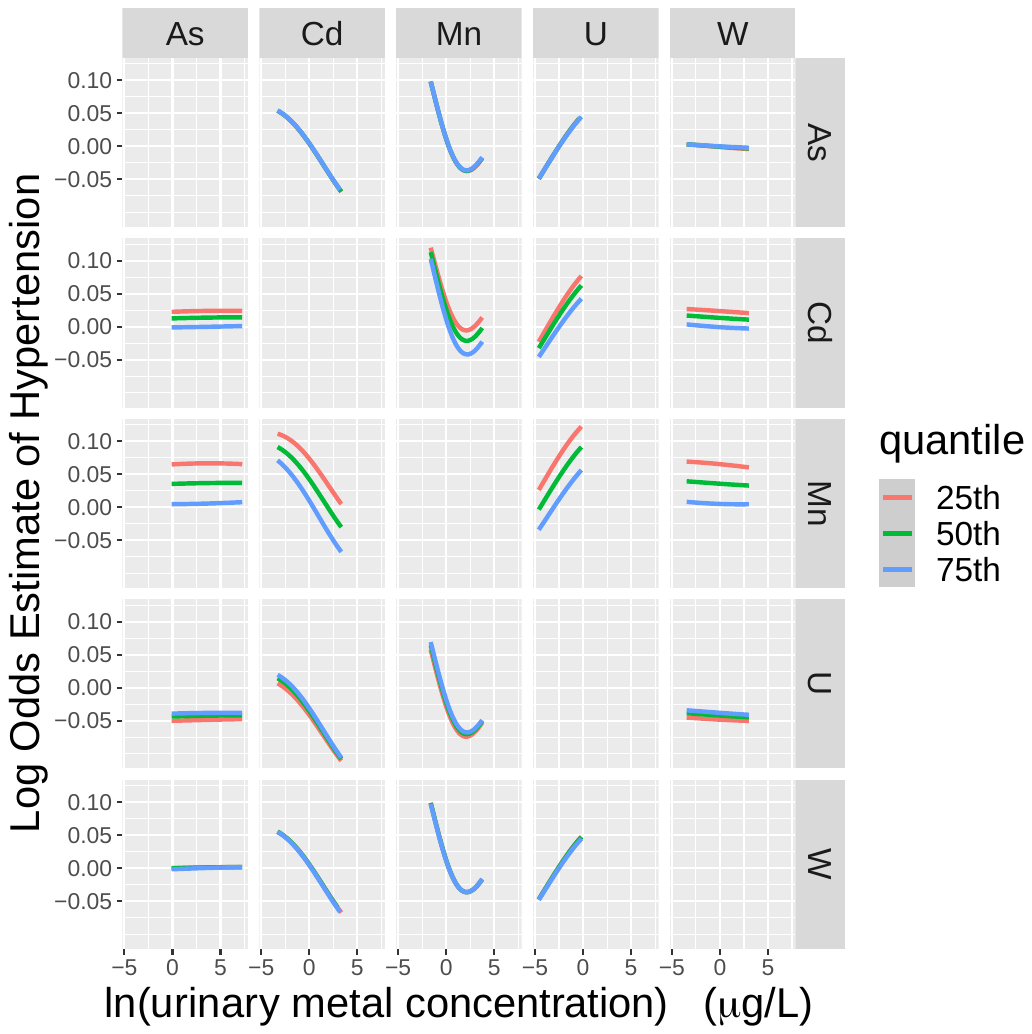

**Supplemental Figure 5C.** Exposure-response functions for the log hazard estimate (interpreted as a log odds estimate) of prevalent hypertension for each metal (columns) when another metal (rows) is held constant at given quantiles and all other metals are held constant at their median. The model was adjusted for sex, age, ethnicity (Hispanic/non-Hispanic), income, obesity (BMI > 30 kg/m^2^), smoking (never/former/current), diet, and urinary creatinine (g/L).

Appendix A: Sources for DAG Arrows

1. Arsenic
   a. As 🡪 CVD (Abdul et al. 2015; Alamolhodaei, Shirani, and Karimi 2015; Boekelheide et al. 2012; Chowdhury et al. 2018; Jomova et al. 2011; Kuo et al. 2017; Leng et al. 2019; Moon et al. 2017; Moon, Guallar, and Navas–Acien 2012; Phung et al. 2017; Solenkova et al. 2014; Stea et al. 2014; Xu, Mondal, and Polya 2020)
   b. As 🡪 diabetes (Abdul et al. 2015; Andra et al. 2013; Beck, Styblo, and Sethupathy 2017; Jomova et al. 2011; Kuo et al. 2013; 2017; Stea et al. 2014; Wang et al. 2014)
   c. As 🡪 hypertension (Abdul et al. 2015; Abhyankar et al. 2012; Jomova et al. 2011; Solenkova et al. 2014; Stea et al. 2014; Xu, Mondal, and Polya 2020)
2. Cadmium
   a. Cd 🡪 CVD (Chowdhury et al. 2018; Diaz et al. 2021; Kukongviriyapan, Apaijit, and Kukongviriyapan 2016; Larsson and Wolk 2016; Satarug, Vesey, and Gobe 2017; Solenkova et al. 2014; Tellez-Plaza et al. 2013; Thévenod and Lee 2013; Tinkov et al. 2018)
   b. Cd 🡪 diabetes (Little et al. 2020; Satarug, Vesey, and Gobe 2017; Thévenod and Lee 2013)
   c. Cd 🡪 hypertension (Caciari et al. 2013; Edwards and Ackerman 2016; Martins et al. 2021; Satarug and Moore 2012; Satarug, Vesey, and Gobe 2017; Solenkova et al. 2014; Thévenod and Lee 2013; Xu, Mondal, and Polya 2020)
3. Manganese
   a. Mn 🡪 CVD (tested in this analysis)
   b. Mn 🡪 diabetes (Kaur and Henry 2014; Li and Yang 2018; Sanjeevi et al. 2018; Siddiqui, Bawazeer, and Scaria Joy 2014)
   c. Mn 🡪 hypertension (tested in this analysis)
4. Uranium
   a. U 🡪 CVD (Nigra et al. 2016)
   b. U 🡪 diabetes (tested in this analysis)
   c. U 🡪 hypertension (tested in this analysis)
5. Tungsten
   a. W 🡪 CVD (Nigra et al. 2016)
   b. W 🡪 diabetes (tested in this analysis)
   c. W 🡪 hypertension (tested in this analysis)
6. Age
   a. Age 🡪 CVD (Wood 2001)
   b. Age 🡪 diabetes (Alberti, Zimmet, and Shaw 2007)
   c. Age 🡪 hypertension (Whelton et al. 2018)
   d. Age 🡪 alcohol (from general knowledge)
   e. Age 🡪 ambient air (Di et al. 2017)
   f. Age 🡪 diet (Winpenny et al. 2018)
   g. Age 🡪 education (from general knowledge)
   h. Age 🡪 physical activity (Telama et al. 2005)
   i. Age 🡪 income (from general knowledge)
7. Alcohol
   a. Alcohol 🡪 CVD (Wood 2001)
   b. Alcohol 🡪 diabetes (Pietraszek, Gregersen, and Hermansen 2010)
   c. Alcohol 🡪 hypertension (Whelton et al. 2018)
8. Ambient air
   a. Ambient air 🡪 As (Balakrishnan et al. 2018; Beck, Styblo, and Sethupathy 2017)
   b. Ambient air 🡪 Cd (Afsar et al. 2019; Bochud et al. 2018; Kukongviriyapan, Apaijit, and Kukongviriyapan 2016; Larsson and Wolk 2016; Satarug, Vesey, and Gobe 2017; Solenkova et al. 2014; Tellez-Plaza et al. 2013)
   c. Ambient air 🡪 Mn (Li and Yang 2018)
   d. Ambient air 🡪 U (Corlin et al. 2016; Nigra et al. 2016)
   e. Ambient air 🡪 CVD (Yang et al. 2019)
   f. Ambient air 🡪 diabetes (Eze et al. 2015)
   g. Ambient air 🡪 hypertension (Cai et al. 2016)
   h. Ambient air 🡪 obesity (Huang et al. 2020)
9. Diet
   a. Diet 🡪 As (Balakrishnan et al. 2018; Moon et al. 2017; Solenkova et al. 2014)
   b. Diet 🡪 Cd (Afsar et al. 2019; Bochud et al. 2018; Edwards and Ackerman 2016; Kukongviriyapan, Apaijit, and Kukongviriyapan 2016; Larsson and Wolk 2016; Satarug, Vesey, and Gobe 2017; Solenkova et al. 2014; Tellez-Plaza et al. 2013)
   c. Diet 🡪 Mn (Li and Yang 2018)
   d. Diet 🡪 U (Corlin et al. 2016)
   e. Diet 🡪 W (Nigra et al. 2016)
   f. Diet 🡪 CVD (Ros et al. 2014; Wood 2001)
   g. Diet 🡪 diabetes (Alberti, Zimmet, and Shaw 2007; Bloomgarden 2002)
   h. Diet 🡪 hypertension (Ozemek et al. 2018; Whelton et al. 2018)
   i. Diet 🡪 obesity (Cheung et al. 2018)
10. Drinking water
    a. Drinking water 🡪 As (Afsar et al. 2019; Balakrishnan et al. 2018; Beck, Styblo, and Sethupathy 2017; Ellinsworth 2015; Kuo et al. 2013; Moon et al. 2017; Solenkova et al. 2014; Tsuji et al. 2014; Wang et al. 2014)
    b. Drinking water 🡪 Cd (Afsar et al. 2019; Solenkova et al. 2014)
    c. Drinking water 🡪 Mn (Li and Yang 2018)
    d. Drinking water 🡪 U (Corlin et al. 2016; Nigra et al. 2016)
    e. Drinking water 🡪 W (Nigra et al. 2016)
11. Education
    a. Education🡪 CVD (Havranek et al. 2015)
    b. Education 🡪 diabetes (Hosseini, Whiting, and Vatanparast 2019; Kim et al. 2017; Seiglie et al. 2020)
    c. Education 🡪 hypertension (Whelton et al. 2018)
    d. Education 🡪 alcohol (Collins 2016)
    e. Education 🡪 diet (Marques-Vidal et al. 2015; C. D. Rehm et al. 2016)
    f. Education 🡪 income (Glomm and Ravikumar 2003)
    g. Education 🡪 physical activity (Zhou and Wang 2019)
    h. Education 🡪 smoking (Huisman, Kunst, and Mackenbach 2005)
12. Ethnicity
    a. Ethnicity 🡪 CVD (Havranek et al. 2015)
    b. Ethnicity 🡪 diabetes (Alberti, Zimmet, and Shaw 2007)
    c. Ethnicity 🡪 hypertension (Whelton et al. 2018)
    d. Ethnicity 🡪 ambient air (Di et al. 2017)
    e. Ethnicity 🡪 drinking water (Schaider et al. 2019)
    f. Ethnicity 🡪 diet (C. D. Rehm et al. 2016; Yau et al. 2020)
    g. Ethnicity 🡪 obesity (Waldstein et al. 2016)
    h. Ethnicity 🡪 smoking (El-Toukhy, Sabado, and Choi 2016)
13. Hypertension
    a. Hypertension 🡪 CVD (Perumareddi 2019; Wood 2001)
14. Income
    a. Income 🡪 CVD (Havranek et al. 2015)
    b. Income 🡪 diabetes (Hosseini, Whiting, and Vatanparast 2019; Seiglie et al. 2020)
    c. Income 🡪 hypertension (Whelton et al. 2018)
    d. Income 🡪 alcohol (Collins 2016)
    e. Income 🡪 ambient air (Di et al. 2017)
    f. Income 🡪 diet (C. D. Rehm et al. 2016)
    g. Income 🡪 drinking water (Schaider et al. 2019)
    h. Income 🡪 obesity (Waldstein et al. 2016; Wardle, Waller, and Jarvis 2002)
    i. Income 🡪 physical activity (SFM et al. 2020)
    j. Income 🡪 smoking (Yun et al. 2015)
15. Obesity
    a. Obesity 🡪 CVD (Wood 2001)
    b. Obesity 🡪 diabetes (Alberti, Zimmet, and Shaw 2007; Maggio and Pi-Sunyer 2003)
    c. Obesity 🡪 hypertension (Seravalle and Grassi 2017; Whelton et al. 2018)
16. Physical activity
    a. Physical activity 🡪 CVD (Havranek et al. 2015; Wood 2001)
    b. Physical activity 🡪 diabetes (Alberti, Zimmet, and Shaw 2007)
    c. Physical activity 🡪 hypertension (Whelton et al. 2018)
    d. Physical activity 🡪 ambient air (Corlin et al. 2019)
    e. Physical activity 🡪 obesity (Swift et al. 2014)
17. Sex
    a. Sex 🡪 CVD (Wood 2001)
    b. Sex 🡪 diabetes (Alberti, Zimmet, and Shaw 2007)
    c. Sex 🡪 hypertension (Whelton et al. 2018)
    d. Sex 🡪 alcohol (J. Rehm et al. 2013)
    e. Sex 🡪 diet (Papier et al. 2015)
    f. Sex 🡪 education (Klasen and Lamanna 2009)
    g. Sex 🡪 income (Klasen and Lamanna 2009)
18. Selection node
    a. Age 🡪 selection (Hamman et al. 1989)
    b. Diabetes 🡪 selection (Hamman et al. 1989)
    c. Ethnicity 🡪 selection (Hamman et al. 1989)
    d. Metal measurement 🡪 selection
    e. Sex 🡪 selection (Hamman et al. 1989)
19. Smoking
    a. Smoking 🡪 Cd (Afsar et al. 2019; Bochud et al. 2018; Edwards and Ackerman 2016; Kukongviriyapan, Apaijit, and Kukongviriyapan 2016; Larsson and Wolk 2016; Satarug, Vesey, and Gobe 2017; Solenkova et al. 2014; Tellez-Plaza et al. 2013)
    b. Smoking 🡪 CVD (Ambrose and Barua 2004; Wood 2001)
    c. Smoking 🡪 diabetes (Maddatu, Anderson-Baucum, and Evans-Molina 2017)
    d. Smoking 🡪 hypertension (Virdis et al. 2010; Whelton et al. 2018)
20. Soil
    a. Soil 🡪 As (Beck, Styblo, and Sethupathy 2017; Solenkova et al. 2014)
    b. Soil 🡪 W (Nigra et al. 2016)
    c. Soil 🡪 diet (Holmgren et al. 1993; Zhuang, Zou, and Shu 2009)
21. Urine creatinine
    a. Urine creatinine 🡪 metal measurement (general knowledge)
    b. Age 🡪 urine creatinine (Barr et al. 2005)
    c. Ethnicity 🡪 urine creatinine (Barr et al. 2005)
    d. Sex 🡪 urine creatinine (Barr et al. 2005)
    e. Obesity 🡪 urine creatinine (Barr et al. 2005)

Appendix B: Minimally Sufficient Adjustment Sets

As, Cd, Mn, W, and U 🡪 cardiovascular disease (CVD)

- Ambient air, diet, drinking water, smoking
- Ambient air, diet, ethnicity, income, smoking

As 🡪 CVD

- Ambient air, Cd, diet, ethnicity, income, Mn, smoking, U, W
- Ambient air, diet, drinking water, soil
- Ambient air, diet, drinking water, W

Cd 🡪 CVD

- Ambient air, As, diet, ethnicity, income, Mn, smoking, U, W
- Ambient air, diet, drinking water, smoking

Mn 🡪 CVD

- Ambient air, As, Cd, diet, ethnicity, income, smoking, U, W
- Ambient air, diet, drinking water

W 🡪 CVD

- Ambient air, As, Cd, diet, ethnicity, income, Mn, smoking, U
- Ambient air, As, diet, drinking water
- Ambient air, diet, drinking water, soil

U 🡪 CVD

- Ambient air, As, Cd, diet, ethnicity, income, Mn, smoking, W
- Ambient air, diet, drinking water

As, Cd, Mn, W, and U 🡪 diabetes

- Ambient air, diet, drinking water, smoking
- Ambient air, diet, ethnicity, income, smoking

As 🡪 diabetes

- Ambient air, Cd, diet, ethnicity, income, Mn, smoking, U, W
- Ambient air, diet, drinking water, soil
- Ambient air, diet, drinking water, W

Cd 🡪 diabetes

- Ambient air, As, diet, ethnicity, income, Mn, smoking, U, W
- Ambient air, diet, drinking water, smoking

Mn 🡪 diabetes

- Ambient air, As, Cd, diet, ethnicity, income, smoking, U, W
- Ambient air, diet, drinking water

W 🡪 diabetes

- Ambient air, As, Cd, diet, ethnicity, income, Mn, smoking, U
- Ambient air, As, diet, drinking water
- Ambient air, diet, drinking water, soil

U 🡪 diabetes

- Ambient air, As, Cd, diet, ethnicity, income, Mn, smoking, W
- Ambient air, diet, drinking water

As, Cd, Mn, W, and U 🡪 hypertension

- Ambient air, diet, drinking water, smoking
- Ambient air, diet, ethnicity, income, smoking

As 🡪 hypertension

- Ambient air, Cd, diet, ethnicity, income, Mn, smoking, U, W
- Ambient air, diet, drinking water, soil
- Ambient air, diet, drinking water, W

Cd 🡪 hypertension

- Ambient air, As, diet, ethnicity, income, Mn, smoking, U, W
- Ambient air, diet, drinking water, smoking

Mn 🡪 hypertension

- Ambient air, As, Cd, diet, ethnicity, income, smoking, U, W
- Ambient air, diet, drinking water

W 🡪 hypertension

- Ambient air, As, Cd, diet, ethnicity, income, Mn, smoking, U
- Ambient air, As, diet, drinking water
- Ambient air, diet, drinking water, soil

U 🡪 hypertension

- Ambient air, As, Cd, diet, ethnicity, income, Mn, smoking, W
- Ambient air, diet, drinking water

DAG Code

dag {

bb="0,0,1,1"

"Ambient air" [pos="0.127,0.572"]

"Drinking water" [pos="0.134,0.721"]

"Metal measurement" [pos="0.151,0.041"]

"Physical activity" [pos="0.639,0.201"]

"Urinary creatinine" [pos="0.146,0.139"]

Age [pos="0.573,0.064"]

Alcohol [pos="0.508,0.814"]

As [pos="0.350,0.137"]

CVD [pos="0.933,0.904"]

Cd [pos="0.352,0.253"]

Diabetes [pos="0.901,0.497"]

Diet [pos="0.080,0.410"]

Education [pos="0.498,0.320"]

Ethnicity [pos="0.693,0.444"]

Hypertension [pos="0.882,0.050"]

Income [pos="0.476,0.601"]

Mn [pos="0.342,0.494"]

Obesity [pos="0.717,0.902"]

Selection [pos="0.398,0.055"]

Sex [pos="0.685,0.613"]

Smoking [pos="0.140,0.910"]

Soil [pos="0.102,0.238"]

U [pos="0.331,0.896"]

W [pos="0.336,0.739"]

"Ambient air" -> As

"Ambient air" -> CVD

"Ambient air" -> Cd

"Ambient air" -> Diabetes

"Ambient air" -> Hypertension

"Ambient air" -> Mn

"Ambient air" -> U

"Ambient air" -> W

"Drinking water" -> As

"Drinking water" -> Cd

"Drinking water" -> Mn

"Drinking water" -> U

"Drinking water" -> W

"Metal measurement" -> Selection

"Physical activity" -> "Ambient air"

"Physical activity" -> CVD

"Physical activity" -> Diabetes

"Physical activity" -> Hypertension

"Physical activity" -> Obesity

"Urinary creatinine" -> "Metal measurement"

Age -> "Ambient air"

Age -> "Physical activity"

Age -> "Urinary creatinine"

Age -> Alcohol

Age -> CVD

Age -> Diabetes

Age -> Diet

Age -> Education

Age -> Hypertension

Age -> Income

Age -> Selection

Alcohol -> CVD

Alcohol -> Diabetes

Alcohol -> Hypertension

As -> CVD

As -> Diabetes

As -> Hypertension

Cd -> CVD

Cd -> Diabetes

Cd -> Hypertension

Diabetes -> Selection

Diet -> As

Diet -> CVD

Diet -> Cd

Diet -> Diabetes

Diet -> Hypertension

Diet -> Mn

Diet -> U

Diet -> W

Education -> "Physical activity"

Education -> Alcohol

Education -> CVD

Education -> Diabetes

Education -> Diet

Education -> Hypertension

Education -> Income

Education -> Smoking

Ethnicity -> "Ambient air"

Ethnicity -> "Drinking water"

Ethnicity -> "Urinary creatinine"

Ethnicity -> CVD

Ethnicity -> Diabetes

Ethnicity -> Diet

Ethnicity -> Hypertension

Ethnicity -> Obesity

Ethnicity -> Selection

Ethnicity -> Smoking

Hypertension -> CVD

Income -> "Ambient air"

Income -> "Drinking water"

Income -> "Physical activity"

Income -> Alcohol

Income -> CVD

Income -> Diabetes

Income -> Diet

Income -> Hypertension

Income -> Obesity

Income -> Smoking

Mn -> CVD

Mn -> Diabetes

Mn -> Hypertension

Obesity -> "Urinary creatinine"

Obesity -> CVD

Obesity -> Diabetes

Obesity -> Hypertension

Sex -> "Urinary creatinine"

Sex -> Alcohol

Sex -> CVD

Sex -> Diabetes

Sex -> Diet

Sex -> Education

Sex -> Hypertension

Sex -> Income

Sex -> Selection

Smoking -> CVD

Smoking -> Cd

Smoking -> Diabetes

Smoking -> Hypertension

Soil -> As

Soil -> W

U -> CVD

U -> Diabetes

U -> Hypertension

W -> CVD

W -> Diabetes

W -> Hypertension

}

Appendix C: Missing Data

| Variable | Number (%) of participants missing data^a^ |
| --- | --- |
| Metals | 186 (10.4) |
| Cardiovascular disease | 135 (7.5) |
| Diabetes | 0 (0) |
| Hypertension | 4 (0.2) |
| Sex | 0 (0) |
| Age | 0 (0) |
| Obesity | 2 (0.1) |
| Education | 190 (10.6) |
| Ethnicity | 0 (0) |
| Income | 136 (7.6) |
| Smoking | 2 (0.1) |
| Physical activity | 8 (0.4) |
| Alcohol | 32 (1.8) |
| Diet | 14 (0.8) |
| Urinary Creatinine | 6 (0.3) |

^a^ Percentages based on total number of participants in SLVDS (n = 1795)

Appendix D: Assessment of Conditional Independence Statements

| Conditional Independence Statement | p-value |
| --- | --- |
| MetalMeasurement ~ PhysicalActivity \| Ethnicity + Obesity + Age + Education + Income | 1.000 |
| MetalMeasurement ~ PhysicalActivity \| Obesity + Age + Sex + Ethnicity | 1.000 |
| MetalMeasurement ~ PhysicalActivity \| UrineCreatinine | 1.000 |
| MetalMeasurement ~ Age \| UrineCreatinine | 1.000 |
| MetalMeasurement ~ Alcohol \| Age + Education + Income + Sex | 1.000 |
| MetalMeasurement ~ Alcohol \| Income + Sex + PhysicalActivity + Age | 1.000 |
| MetalMeasurement ~ Alcohol \| Ethnicity + Obesity + Age + Sex | 1.000 |
| MetalMeasurement ~ Alcohol \| UrineCreatinine | 1.000 |
| MetalMeasurement ~ As \| Ethnicity + Income + PhysicalActivity + Age + Diet1 + Diet2 | 1.000 |
| MetalMeasurement ~ As \| Ethnicity + Income + Sex + PhysicalActivity + Age | 1.000 |
| MetalMeasurement ~ As \| Age + Ethnicity + Income + Obesity + Education + Diet1 + Diet2 | 1.000 |
| MetalMeasurement ~ As \| Age + Ethnicity + Obesity + Sex | 1.000 |
| MetalMeasurement ~ As \| UrineCreatinine | 1.000 |
| MetalMeasurement ~ CVD \| Ethnicity + Age + Obesity + Sex | 1.000 |
| MetalMeasurement ~ CVD \| UrineCreatinine | 1.000 |
| MetalMeasurement ~ Cd \| SmokingStatus + Ethnicity + Income + PhysicalActivity + Age + Diet1 + Diet2 | 1.000 |
| MetalMeasurement ~ Cd \| Education + Ethnicity + Income + PhysicalActivity + Age + Diet1 + Diet2 | 1.000 |
| MetalMeasurement ~ Cd \| Ethnicity + Income + Sex + PhysicalActivity + Age | 1.000 |
| MetalMeasurement ~ Cd \| Age + Ethnicity + Income + Obesity + Education + Diet1 + Diet2 | 1.000 |
| MetalMeasurement ~ Cd \| Age + Ethnicity + Obesity + Sex | 1.000 |
| MetalMeasurement ~ Cd \| UrineCreatinine | 1.000 |
| MetalMeasurement ~ Diabetes \| Ethnicity + Age + Obesity + Sex | 1.000 |
| MetalMeasurement ~ Diabetes \| UrineCreatinine | 1.000 |
| MetalMeasurement ~ Diet1 \| Age + Education + Ethnicity + Income + Sex | 1.000 |
| MetalMeasurement ~ Diet1 \| Ethnicity + Income + Sex + PhysicalActivity + Age | 1.000 |
| MetalMeasurement ~ Diet1 \| Ethnicity + Obesity + Age + Sex | 1.000 |
| MetalMeasurement ~ Diet1 \| UrineCreatinine | 1.000 |
| MetalMeasurement ~ Education \| Income + PhysicalActivity + Age + Sex | 1.000 |
| MetalMeasurement ~ Education \| Ethnicity + Obesity + Age + Sex | 1.000 |
| MetalMeasurement ~ Education \| UrineCreatinine | 1.000 |
| MetalMeasurement ~ Ethnicity \| UrineCreatinine | 1.000 |
| MetalMeasurement ~ Hypertension \| Ethnicity + Age + Obesity + Sex | 1.000 |
| MetalMeasurement ~ Hypertension \| UrineCreatinine | 1.000 |
| MetalMeasurement ~ Income \| Ethnicity + Obesity + Age + Sex | 1.000 |
| MetalMeasurement ~ Income \| UrineCreatinine | 1.000 |
| MetalMeasurement ~ Mn \| Ethnicity + Income + PhysicalActivity + Age + Diet1 + Diet2 | 1.000 |
| MetalMeasurement ~ Mn \| Ethnicity + Income + Sex + PhysicalActivity + Age | 1.000 |
| MetalMeasurement ~ Mn \| Age + Ethnicity + Income + Obesity + Education + Diet1 + Diet2 | 1.000 |
| MetalMeasurement ~ Mn \| Age + Ethnicity + Obesity + Sex | 1.000 |
| MetalMeasurement ~ Mn \| UrineCreatinine | 1.000 |
| MetalMeasurement ~ Obesity \| UrineCreatinine | 1.000 |
| MetalMeasurement ~ Sex \| UrineCreatinine | 1.000 |
| MetalMeasurement ~ SmokingStatus \| Education + Ethnicity + Income | 1.000 |
| MetalMeasurement ~ SmokingStatus \| Ethnicity + Income + Sex + PhysicalActivity + Age | 1.000 |
| MetalMeasurement ~ SmokingStatus \| Ethnicity + Obesity + Age + Sex | 1.000 |
| MetalMeasurement ~ SmokingStatus \| UrineCreatinine | 1.000 |
| MetalMeasurement ~ U \| Ethnicity + Income + PhysicalActivity + Age + Diet1 + Diet2 | 1.000 |
| MetalMeasurement ~ U \| Ethnicity + Income + Sex + PhysicalActivity + Age | 1.000 |
| MetalMeasurement ~ U \| Age + Ethnicity + Income + Obesity + Education + Diet1 + Diet2 | 1.000 |
| MetalMeasurement ~ U \| Age + Ethnicity + Obesity + Sex | 1.000 |
| MetalMeasurement ~ U \| UrineCreatinine | 1.000 |
| MetalMeasurement ~ W \| Ethnicity + Income + PhysicalActivity + Age + Diet1 + Diet2 | 1.000 |
| MetalMeasurement ~ W \| Ethnicity + Income + Sex + PhysicalActivity + Age | 1.000 |
| MetalMeasurement ~ W \| Age + Ethnicity + Income + Obesity + Education + Diet1 + Diet2 | 1.000 |
| MetalMeasurement ~ W \| Age + Ethnicity + Obesity + Sex | 1.000 |
| MetalMeasurement ~ W \| UrineCreatinine | 1.000 |
| PhysicalActivity ~ UrineCreatinine \| Age + Ethnicity + Obesity + Sex | 0.392 |
| PhysicalActivity ~ UrineCreatinine \| Income + Age + Ethnicity + Obesity + Education | 0.414 |
| PhysicalActivity ~ Alcohol \| Age + Education + Income | 0.086 |
| PhysicalActivity ~ Diet1 \| Age + Education + Income | <0.001 |
| PhysicalActivity ~ Ethnicity | <0.001 |
| PhysicalActivity ~ Sex \| Income + Age + Education | <0.001 |
| PhysicalActivity ~ SmokingStatus \| Income + Education | 0.613 |
| UrineCreatinine ~ Alcohol \| Age + Education + Income + Sex | 0.296 |
| UrineCreatinine ~ Alcohol \| Income + Sex + PhysicalActivity + Age | 0.521 |
| UrineCreatinine ~ Alcohol \| Ethnicity + Obesity + Age + Sex | 0.336 |
| UrineCreatinine ~ As \| Ethnicity + Income + PhysicalActivity + Age + Diet1 + Diet2 | <0.001 |
| UrineCreatinine ~ As \| Ethnicity + Income + Sex + PhysicalActivity + Age | <0.001 |
| UrineCreatinine ~ As \| Age + Ethnicity + Income + Obesity + Education + Diet1 + Diet2 | <0.001 |
| UrineCreatinine ~ As \| Age + Ethnicity + Obesity + Sex | <0.001 |
| UrineCreatinine ~ CVD \| Ethnicity + Age + Obesity + Sex | 0.222 |
| UrineCreatinine ~ Cd \| SmokingStatus + Ethnicity + Income + PhysicalActivity + Age + Diet1 + Diet2 | <0.001 |
| UrineCreatinine ~ Cd \| Education + Ethnicity + Income + PhysicalActivity + Age + Diet1 + Diet2 | <0.001 |
| UrineCreatinine ~ Cd \| Ethnicity + Income + Sex + PhysicalActivity + Age | <0.001 |
| UrineCreatinine ~ Cd \| Age + Ethnicity + Income + Obesity + Education + Diet1 + Diet2 | <0.001 |
| UrineCreatinine ~ Cd \| Age + Ethnicity + Obesity + Sex | <0.001 |
| UrineCreatinine ~ Diabetes \| Ethnicity + Age + Obesity + Sex | <0.001 |
| UrineCreatinine ~ Diet1 \| Age + Education + Ethnicity + Income + Sex | 0.401 |
| UrineCreatinine ~ Diet1 \| Ethnicity + Income + Sex + PhysicalActivity + Age | 0.192 |
| UrineCreatinine ~ Diet1 \| Ethnicity + Obesity + Age + Sex | 0.389 |
| UrineCreatinine ~ Education \| Income + PhysicalActivity + Age + Sex | 0.149 |
| UrineCreatinine ~ Education \| Ethnicity + Obesity + Age + Sex | 0.006 |
| UrineCreatinine ~ Hypertension \| Ethnicity + Age + Obesity + Sex | <0.001 |
| UrineCreatinine ~ Income \| Ethnicity + Obesity + Age + Sex | 0.019 |
| UrineCreatinine ~ Mn \| Ethnicity + Income + PhysicalActivity + Age + Diet1 + Diet2 | <0.001 |
| UrineCreatinine ~ Mn \| Ethnicity + Income + Sex + PhysicalActivity + Age | <0.001 |
| UrineCreatinine ~ Mn \| Age + Ethnicity + Income + Obesity + Education + Diet1 + Diet2 | <0.001 |
| UrineCreatinine ~ Mn \| Age + Ethnicity + Obesity + Sex | <0.001 |
| UrineCreatinine ~ SmokingStatus \| Education + Ethnicity + Income | 0.008 |
| UrineCreatinine ~ SmokingStatus \| Ethnicity + Income + Sex + PhysicalActivity + Age | 0.406 |
| UrineCreatinine ~ SmokingStatus \| Ethnicity + Obesity + Age + Sex | 0.379 |
| UrineCreatinine ~ U \| Ethnicity + Income + PhysicalActivity + Age + Diet1 + Diet2 | <0.001 |
| UrineCreatinine ~ U \| Ethnicity + Income + Sex + PhysicalActivity + Age | <0.001 |
| UrineCreatinine ~ U \| Age + Ethnicity + Income + Obesity + Education + Diet1 + Diet2 | <0.001 |
| UrineCreatinine ~ U \| Age + Ethnicity + Obesity + Sex | <0.001 |
| UrineCreatinine ~ W \| Ethnicity + Income + PhysicalActivity + Age + Diet1 + Diet2 | <0.001 |
| UrineCreatinine ~ W \| Ethnicity + Income + Sex + PhysicalActivity + Age | <0.001 |
| UrineCreatinine ~ W \| Age + Ethnicity + Income + Obesity + Education + Diet1 + Diet2 | <0.001 |
| UrineCreatinine ~ W \| Age + Ethnicity + Obesity + Sex | <0.001 |
| Age ~ Ethnicity | 0.726 |
| Age ~ Obesity \| Income + PhysicalActivity | 0.382 |
| Age ~ Sex | 0.914 |
| Age ~ SmokingStatus \| Income + Education | 0.077 |
| Alcohol ~ As \| Ethnicity + Income + PhysicalActivity + Age + Diet1 + Diet2 | <0.001 |
| Alcohol ~ As \| Age + Ethnicity + Income + Education + Diet1 + Diet2 | <0.001 |
| Alcohol ~ As \| Age + Income + Education + Sex | <0.001 |
| Alcohol ~ Cd \| SmokingStatus + Ethnicity + Income + PhysicalActivity + Age + Diet1 + Diet2 | 0.061 |
| Alcohol ~ Cd \| Age + Ethnicity + Income + Education + Diet1 + Diet2 | 0.024 |
| Alcohol ~ Cd \| Age + Income + Education + Sex | 0.015 |
| Alcohol ~ Diet1 \| Income + Sex + Age + Education | 0.038 |
| Alcohol ~ Ethnicity | 0.450 |
| Alcohol ~ Mn \| Ethnicity + Income + PhysicalActivity + Age + Diet1 + Diet2 | 0.954 |
| Alcohol ~ Mn \| Age + Ethnicity + Income + Education + Diet1 + Diet2 | 0.949 |
| Alcohol ~ Mn \| Age + Income + Education + Sex | 0.979 |
| Alcohol ~ Obesity \| Income + PhysicalActivity | 0.020 |
| Alcohol ~ Obesity \| Income + Age + Education | 0.019 |
| Alcohol ~ SmokingStatus \| Income + Education | <0.001 |
| Alcohol ~ U \| Ethnicity + Income + PhysicalActivity + Age + Diet1 + Diet2 | 0.093 |
| Alcohol ~ U \| Age + Ethnicity + Income + Education + Diet1 + Diet2 | 0.091 |
| Alcohol ~ U \| Age + Income + Education + Sex | 0.142 |
| Alcohol ~ W \| Ethnicity + Income + PhysicalActivity + Age + Diet1 + Diet2 | <0.001 |
| Alcohol ~ W \| Age + Ethnicity + Income + Education + Diet1 + Diet2 | <0.001 |
| Alcohol ~ W \| Age + Income + Education + Sex | <0.001 |
| As ~ Education \| Age + Ethnicity + Income + PhysicalActivity + Diet1 + Diet2 | 0.553 |
| As ~ Obesity \| PhysicalActivity + Ethnicity + Income | 0.746 |
| As ~ Sex \| Age + Education + Ethnicity + Income + Diet1 + Diet2 | 0.058 |
| As ~ Sex \| Ethnicity + Income + PhysicalActivity + Age + Diet1 + Diet2 | 0.052 |
| As ~ SmokingStatus \| Education + Ethnicity + Income | 0.005 |
| As ~ SmokingStatus \| Income + Age + Ethnicity + PhysicalActivity + Diet1 + Diet2 | 0.004 |
| Cd ~ Education \| Income + Age + Ethnicity + SmokingStatus + PhysicalActivity + Diet1 + Diet2 | 0.537 |
| Cd ~ Obesity \| PhysicalActivity + Ethnicity + Income | 0.160 |
| Cd ~ Sex \| Income + Age + Education + Ethnicity + Diet1 + Diet2 | 0.610 |
| Cd ~ Sex \| Ethnicity + Income + SmokingStatus + PhysicalActivity + Age + Diet1 + Diet2 | 0.685 |
| Diet1 ~ Obesity \| PhysicalActivity + Ethnicity + Income | 0.041 |
| Diet1 ~ Obesity \| Ethnicity + Income + Age + Education | 0.018 |
| Diet1 ~ SmokingStatus \| Education + Ethnicity + Income | 0.082 |
| Education ~ Ethnicity | <0.001 |
| Education ~ Mn \| Ethnicity + Income + PhysicalActivity + Age + Diet1 + Diet2 | 0.547 |
| Education ~ Obesity \| Income + PhysicalActivity | 0.070 |
| Education ~ U \| Ethnicity + Income + PhysicalActivity + Age + Diet1 + Diet2 | 0.717 |
| Education ~ W \| Ethnicity + Income + PhysicalActivity + Age + Diet1 + Diet2 | 0.209 |
| Ethnicity ~ Income | <0.001 |
| Ethnicity ~ Sex | 0.101 |
| Mn ~ Obesity \| PhysicalActivity + Ethnicity + Income | 0.114 |
| Mn ~ Sex \| Age + Education + Ethnicity + Income + Diet1 + Diet2 | 0.362 |
| Mn ~ Sex \| Ethnicity + Income + PhysicalActivity + Age + Diet1 + Diet2 | 0.388 |
| Mn ~ SmokingStatus \| Education + Ethnicity + Income | 0.022 |
| Mn ~ SmokingStatus \| Income + Age + Ethnicity + PhysicalActivity + Diet1 + Diet2 | 0.010 |
| Obesity ~ Sex \| Income + Age + Education | 0.002 |
| Obesity ~ Sex \| Income + PhysicalActivity | 0.006 |
| Obesity ~ SmokingStatus \| Education + Ethnicity + Income | <0.001 |
| Obesity ~ SmokingStatus \| Income + PhysicalActivity + Ethnicity | <0.001 |
| Obesity ~ U \| Ethnicity + Income + PhysicalActivity | 0.071 |
| Obesity ~ W \| Ethnicity + Income + PhysicalActivity | 0.679 |
| Sex ~ SmokingStatus \| Income + Education | <0.001 |
| Sex ~ U \| Ethnicity + Income + PhysicalActivity + Age + Diet1 + Diet2 | 0.093 |
| Sex ~ U \| Age + Ethnicity + Income + Education + Diet1 + Diet2 | 0.080 |
| Sex ~ W \| Ethnicity + Income + PhysicalActivity + Age + Diet1 + Diet2 | 0.014 |
| Sex ~ W \| Age + Ethnicity + Income + Education + Diet1 + Diet2 | 0.008 |
| SmokingStatus ~ U \| Ethnicity + Income + PhysicalActivity + Age + Diet1 + Diet2 | 0.009 |
| SmokingStatus ~ U \| Ethnicity + Income + Education | 0.006 |
| SmokingStatus ~ W \| Ethnicity + Income + PhysicalActivity + Age + Diet1 + Diet2 | 0.130 |
| SmokingStatus ~ W \| Ethnicity + Income + Education | 0.119 |
